# A genome-wide deletion map in 125,730 individuals for novel rare disease gene and variant discovery

**DOI:** 10.64898/2026.05.13.26352722

**Authors:** Anthony McGuigan, Alistair T. Pagnamenta, Laura E. Covill, Jacob Samson, Carme Camps, Yuyang Chen, Tanishi Moitra, Kartik Chundru, Emily O’Heir, Kirsten Allan, Gavin Arno, Alexander Broomfield, Martin Delatycki, Siying Lin, Michel Michaelides, Rocio Rius, Tony Roscioli, Cas Simons, Andrew Webster, Susan M. White, Louise Wilson, Stephan J. Sanders, Anne O’Donnell-Luria, Jamie M. Ellingford, Jenny C. Taylor, Nicola Whiffin

**Affiliations:** Centre for Human Genetics, NIHR Oxford Biomedical Research Centre, University of Oxford, Oxford, UK; Big Data Institute, University of Oxford, Oxford, UK; Department of Clinical and Biomedical Sciences, Faculty of Health and Life Sciences, University of Exeter, Exeter, UK; Program in Medical and Population Genetics, Broad Institute of MIT and Harvard, Cambridge, MA, USA; Manton Center for Orphan Disease Research, Division of Genetics and Genomics, Boston Children’s Hospital, Harvard Medical School, Boston, MA, USA; Division of Evolution, Infection and Genomics, School of Biological Sciences, Faculty of Biology, Medicine and Health, University of Manchester, Manchester, UK; Department of Life Sciences, Imperial College London, London, UK; Department of Genomic Medicine, University of Cambridge, Cambridge, UK; Victorian Clinical Genetics Services, Murdoch Children’s Research Institute, Melbourne, Australia; NIHR Biomedical Research Centre, Moorfields Eye Hospital and the UCL Institute of Ophthalmology, London, UK; Greenwood Genetic Center, Greenwood, South Carolina, USA; Metabolic Department, Great Ormond Street Hospital, London, UK; Department of Paediatrics, University of Melbourne, Melbourne, Australia; Manchester Centre for Genomic Medicine, Saint Mary’s Hospital, Manchester University NHS Foundation Trust, Manchester, UK; Centre for Population Genomics, Garvan Institute of Medical Research, and UNSW Sydney, Sydney, Australia; Centre for Population Genomics, Murdoch Children’s Research Institute, Melbourne, Australia; Neuroscience Research Australia (NeuRA), Sydney, Australia; Prince of Wales Clinical School, University of New South Wales, Sydney, Australia; NSW Health Pathology Randwick Genomics, Randwick, Australia; Clinical Genetics, North East Thames Regional Genetics Service, Great Ormond Street Hospital for Children NHS Foundation Trust, London, UK; Institute of Developmental and Regenerative Medicine, Department of Paediatrics, University of Oxford, Oxford, UK; Department of Psychiatry and Behavioral Sciences, UCSF Weill Institute for Neurosciences, University of California, San Francisco, San Francisco, CA, USA; New York Genome Center, New York, USA

## Abstract

Structural variants (SVs) can disrupt gene function and contribute to pathogenesis of rare disorders. Here, we created a genome-wide knockout dataset across 125,730 individuals with genome sequencing data in the UK’s National Genomic Research Library by leveraging the distinct read-depth signal associated with homozygous deletions. We curated 535,699 rare high-confidence homozygous deletion SVs, of which 48,735 were rare. These deletions collectively covered 213Mb or 6.92% of the human genome (4.58% of autosomal sequence), revealing substantial tolerance to complete sequence loss. From a subset of 58,022 individuals with rare disease, we identified 295 individuals with likely diagnostic homozygous deletions impacting protein-coding regions of known disease genes. A further 32 individuals had candidate non-coding SVs in or near to known disease genes, 19/32 (59.37%) of which disrupted 5’-UTR/promoter regions, revealing promoter deletion as an underappreciated cause of rare disorders. Finally, we identify 43 genes with no known rare-disease association but with exonic homozygous deletions in two or more individuals with consistent phenotypes. We describe in detail *PDC* (phosducin) in Leber Congenital Amaurosis, *GCG* (glucagon) for a syndromic neurodevelopmental disorder with gastrointestinal involvement, and *ENTPD3* for intellectual disability with autism, as candidate novel disease-associated genes. Overall, we create a genome-wide map of homozygous deletions and demonstrate the power of this dataset for rare disease diagnosis and novel disease-gene discovery.

## Main

Rare disorders collectively impact over 300 million people worldwide^1^. Over 80% of rare disorders are thought to have a genetic cause and identifying that cause can significantly improve familial and clinical management, including access to treatment^1–3^. Despite advances in genome sequencing technologies, around half of all rare disease patients remain without a genetic diagnosis^4,5^. Recently, progress in detection of structural variants (SVs) in short-read exome and genome sequencing data has demonstrated that they are a significant contributor to previously missed rare disease diagnoses^6–8^.

Structural variants (SVs) represent a diverse group of genomic alterations, encompassing deletions, duplications, insertions, inversions, translocations, and complex combinations that may occur concurrently within a single mutational event^9^. SVs are implicated in a diverse spectrum of genetic disorders; germline SVs are a frequent cause of rare disease^10,11^, while somatic SVs (e.g. translocations creating a pathogenic fusion gene) are a common driver event in some cancers^12,13^. Whether germline or somatic, many SVs likely cause disease by altering the baseline gene dosage above or below a tolerated threshold^14^. This can be achieved directly, if an SV overlaps a gene, or indirectly, such as through disruption of critical regulatory elements or 3D genomic organisation. This latter group of non-coding SVs that act via disrupting native gene expression levels^15–17^, are less well characterised compared to coding SVs. Demonstrative examples include promoter deletions reducing cognate gene expression^18^ and deletions within topologically associated domains (TADs) that cause ectopic gene expression through aberrant enhancer activity^19^.

SVs contribute significantly to genomic diversity and human disease through a variety of molecular mechanisms, but their accurate identification remains challenging. Although SV detection has improved with long-read sequencing, the vast majority of clinical genetic testing uses short-read sequencing technologies, which have inherent limitations in SV detection: true SVs are frequently missed, while many of the variants that are identified turn out to be false positives^20^. The signal-to-noise ratio for SV calling varies considerably depending on the calling software, SV type, SV size, and the surrounding sequence context, particularly in repetitive or GC rich genomic regions.

False positive rates vary substantially (e.g. 4%-91% for deletions^21,22^) with increased specificity corresponding to a large drop in sensitivity. In the context of rare disease diagnosis, where high sensitivity is often desired, the resulting high false positive rate makes manual and/or experimental confirmation of all identified SVs infeasible as each individual harbors more than 20,000 putative SVs^23^. Outside of large-scale SVs detectable by karyotyping, the most straightforward SV type to identify from short read genome sequencing are homozygous deletions: while certain features, such as split reads at breakpoints, are shared among various SV types, a complete absence of mapped reads across a normally well-covered region is a distinctive hallmark of homozygous deletions.

Mapping the landscape of deletions and duplication SVs (i.e. copy number variants; CNVs) across the genome provides context to genomic regions where a change in amount of DNA may lead to disease or otherwise may likely be tolerated. In the latter case, deletion SVs can represent ‘natural human knockouts’ of genes or genomic regions which are an informative biomarker for therapeutic knockdown or inhibition. This concept has been investigated through homozygous predicted loss-of-function (pLoF) variants within protein-coding genes^24–26^. Previous studies have also extensively mapped heterozygous CNVs in human cohorts to study changes in gene dosage^27^.

However, despite their added value, tolerance to homozygous deletion SVs has been less well studied on a large scale. Here, we develop a computational approach that leverages read-depth signals to curate a high-confidence set of homozygous deletions across 125,730 participants in the UK’s

National Genome Research Library^28^ (NGRL). We use these data to create a genome-wide map of tolerance to homozygous deletion, showing that >8% of the genome is tolerant of complete loss. Through intersecting genomic deletions identified in rare disease patients with rich phenotypic data within the NGRL, we uncover new diagnoses across a broad range of disorders and identify novel candidate gene-disease relationships.

## Results

### A significant proportion of the genome is tolerant of homozygous deletion

We obtained putative biallelic deletions identified by Canvas^29^, DRAGEN^30^ and Manta^31^ in 125,730 individuals from the NGRL, comprising participants from both the Genomics England 100,000 Genomes Project^32^ (100kGP) and the UK National Health Service Genomic Medicine Service^33^ (NHS-GMS). Of the individuals, 58,022 (46.3%) were affected with a rare disorder, 50,484 (40.3%) were unaffected family members of rare disease probands, and 17,224 (13.4%) were participants recruited with cancer. The homozygous deletions were aggregated into a unified SV set where those with >80% overlap were considered the same event. High-confidence deletions were defined as those with median read-depth ≤1 in the participant and ≥10 across a sample of control individuals. These thresholds were chosen based on the coverage depth across 34 biallelic deletions which were previously confirmed molecular diagnoses; see **Methods**; **Supplemental Fig. 1**).

In the 125,730 participants, we identified 64,585,214 high-confidence homozygous deletion SVs, corresponding to a median of 464 biallelic deletions per individual, with variation depending on cohort (**Fig. 1**). This compares to a median of 1,781 homozygous deletion SVs identified per individual without our read-depth filtering, demonstrating a large reduction in noise across our high confidence set (**Supplemental Fig. 2)**. Of these, 5,377,394 homozygous deletions (8.33%) were rare (1,350,375 unique SVs; cohort AF ≤ 0.001; median 8 per participant; **Supplemental Fig. 3**). While the density of deletion breakpoints was enriched around regions of low genomic complexity, the requirement for a minimum read depth across the deletion in controls mitigated against spurious calls such as in the centromeres as shown for chromosome 1 (**Fig. 1C**).

**Figure 1:**
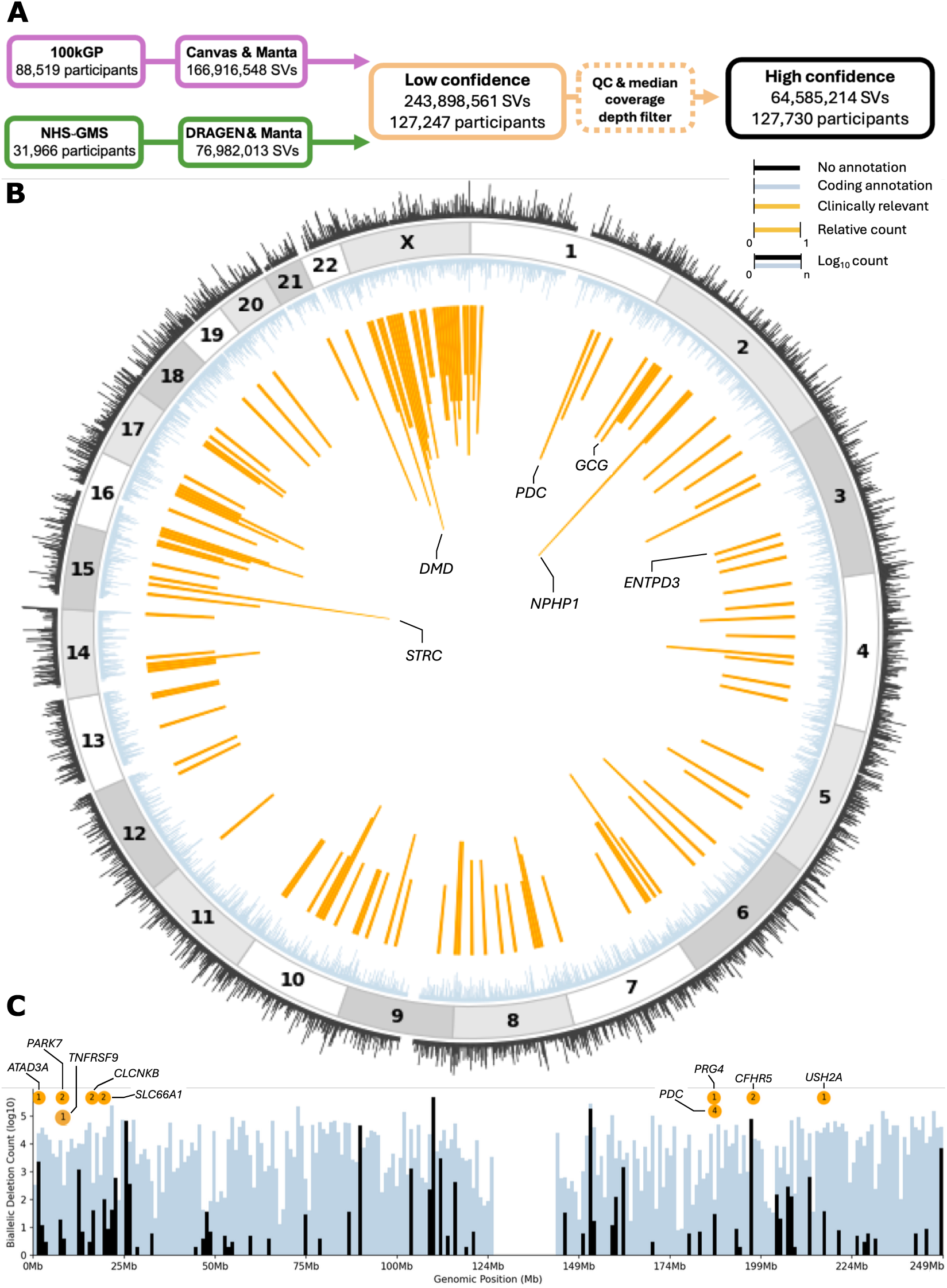
High confidence biallelic deletion dataset. **(A)** Flow diagram of participant and variant processing pipeline. Variants from GRCh37 aligned participants (209,888 unique variants in 7,843 individuals) were lifted over to GRCh38 after depth filtering. **(B)** Circos plot mapping the spectrum of unique genomic sequence deleted (1 megabase bins) by high-confidence SVs in our set. Coding regions and diagnostic deletions highlighted. **(C)** Linear representation of chr1 SV data as seen in (B), log normalized. Diagnostic SVs highlighted in orange with the number of diagnoses enclosed within the circle.

We observed similar numbers of biallelic deletions across male and females, with the exception of chromosome X (**Supplemental Fig. 4**), where hemizygous deletions identified in males affect the ratio between sexes.

The identified deletions ranged in size from 50bp to 6.28Mb with a median length of 264bp across all SV occurrences, or 3.80Kb counting each unique SV once. A characteristic peak was seen at ∼300bp due to polymorphic Alu elements present in the reference genome being called as homozygous deletions when the Alu element is missing in a participant^34^ (**Supplemental Fig. 5**).

A mean of 0.025% or 77Mb of the genome was deleted per individual (range 0.01-0.62%). Across all individuals, homozygous deletions covered 213 million unique nucleotides, corresponding to 6.92% of the GRCh38 human genome (**Supplemental Fig. 2**). The X chromosome accounted for a disproportionate amount of our ‘knockout’ deletions; 52.6% of the chromosome sequence was covered by deletions in our high confidence set, driven by hemizygous deletions identified in males. Chromosome Y had the lowest chromosomal coverage at 0.5%, where low mapping quality meant very few variants passed the minimum depth requirement across controls for SV inclusion. Considering only the autosomal sequence, 5.82% of bases were deleted in at least one individual (**Supplemental Fig. 2**).

As expected, tolerance to homozygous deletion varied by genome annotation, with only 3.11% of protein-coding bases deleted in at least one individual compared to 4.74% of non-coding exonic bases, 5.81% of intronic bases, and 9.41% of intergenic sequence (**Supplemental Table 1**). Of the 1,361,635 unique deletions we identified, 427,266 (31.4%) overlapped protein-coding genes (**Supplemental Table 2**). Only a minority of these variants were predicted to truncate the gene’s coding sequence (63,023/427,266; 14.75%). Even fewer (506/427,266; 0.12%) were predicted to impact the protein-coding sequence of a gene with a known disease annotation (see **Methods**), and individuals with these variants were much more likely to be affected by a rare disease compared to individuals without such variants (303/384 versus 60,402/125,346; OR = 4.66, Chi-Square test, P = 4.86×10^-38^).

### 0.5% of rare disease families have diagnoses due to knockout of known disease genes

#### Protein coding deletions

Of the 63,023 unique homozygous deletions predicted to truncate the coding sequence of a protein-coding region (**Supplemental Table 2**), 506 (identified in 959 individuals) impacted known biallelic disease-associated genes (PanelApp^35^ and Online Mendelian Inheritance in Man^36^ (OMIM)). After manual review (see **Methods**), 303 deletion events across 295 individuals were flagged as likely diagnoses (**Supplemental Table 3**). This corresponded to 0.5% of all individuals (n=58,458) recruited for a rare disease. The likely diagnostic deletions were found across 83 recruited diseases (**Supplemental Fig. 6**). While the most common phenotype in the cohort was Intellectual Disability/Neurodevelopmental Delay (ID/NDD; 16.35% of diagnoses), there was a depletion of ID/NDD individuals in our biallelic SV diagnoses compared to the proportion of individuals in the NGRL with ID/NDD (biallelic SV 46/312, NGRL 2,309/10,797; Chi-Square test, OR = 0.62; *P* = 3.90×10^-3^). This is consistent with these disorders being predominantly driven by *de novo* variants^37,38^.

A large proportion of the likely diagnostic protein-coding deletions (108/303; 35.64%) were affecting genes disrupted in a single proband, however many diagnoses resulted from recurrent SVs, including known deletion-prone loci in *STRC*^39^ and *OTOA*^40^ leading to hearing loss, along with founder variants such as in *NPHP1*^41^ in nephronophthisis **(Fig. 2A)**.

**Figure 2:**
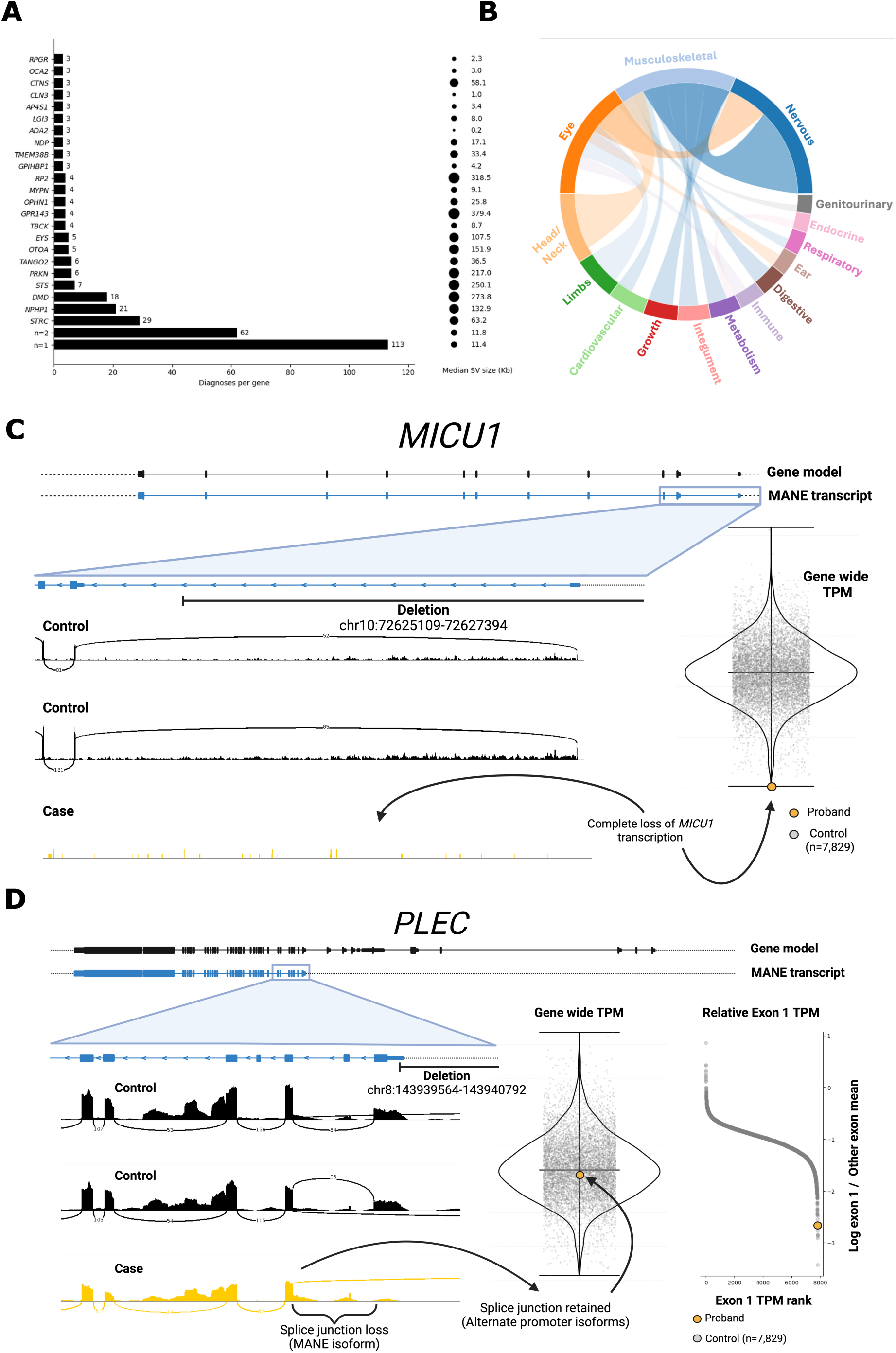
Diagnoses in known disease genes. **(A)** Diagnosis count per gene, ranked by number of occurrences. Includes protein-coding (n=303) and non-coding (n=19) diagnoses across the known disease gene set. n=2 or n=1, genes are binned into a single group. The median SV size of the diagnostic variant is given in kilobases, rounded to one decimal place. **(B)** Circos plot of HPO terms seen across the diagnoses. HPOs were filtered to the ‘system’ level in the HPO hierarchy. HPO text string ‘Abnormality of [the] [X]’ truncated to just [X], where X is the system exhibiting a disease phenotype. HPO connections are displayed in the centre of the circos plot. Where an individual proband had multiple HPOs, they were filtered to show the two most common connections per HPO. **(C)** Schematic of *MICU1*, deleting the first non-coding exon of the 5’UTR and 2kb of upstream sequence. Blood RNA-sequencing from the proband was compared to all transcriptomics samples (n=7,829), showing that the proband had the single lowest *MICU1* expression across all samples. **(D)** Schematic of *PLEC* SV, deleting 20bp of 5’ UTR and a further 1.1kb upstream. Blood RNA-sequencing data from the proband was compared to all transcriptomics samples (n=7,829): the proband has normal *PLEC* expression (across all blood expressed transcripts), however, has loss of the splice junction between exon 1 and 2 (MANE Select transcript). Log ratio of the mean expression of all other *PLEC* exons vs exon 1 shows this proband has the 6^th^ most skewed expression (7,824/7,829) against exon 1, suggesting a transcript specific effect.

Of the 295 individuals with protein-coding deletions, the deletion was recorded in the NGRL as the diagnostic variant for 87/295 (29.49%). The other 208 have now been reported through the Genomics England Diagnostic Discovery pipeline, a subset of which were already known to the internal team. Our approach successfully identified all previously known homozygous deletion diagnoses. For three individuals, the SV was an additional diagnosis in a partially solved case. Six probands had a large biallelic SV overlapping the coding sequence of multiple disease-associated genes.

Probands with putative diagnostic homozygous deletions were almost nine-fold more likely to have consanguineous parents compared to the average across 100kGP; of 30,655 probands where consanguinity status was recorded, 64/177 (36.16%) were in our list of probands with a clinically relevant biallelic deletion compared to 1,826/28,652 (6.37%) other 100kGP probands (Fisher’s exact test, OR = 8.89, *P* = 1.72×10^-32^).

### Non-coding promoter deletions are an under-recognised cause of rare disorders

Deletions that truncate the protein-coding sequence are a well-established cause of gene dysfunction and disease, but non-coding deletions are not often systematically assessed, at least outside of well-studied loci. We first searched for deletions impacting non-coding regions of protein-coding genes. Specifically, we searched for deletions overlapping either the 5’UTR or 3’UTR, and/or the region immediately upstream of the transcript start site (overlapping an ENCODE^42^ candidate promoter within 2kb of the disease-associated gene), which also did not overlap with the coding sequence. We identified 32 such variants across disease-associated protein-coding genes in the context of recessive loss-of-function. Manual curation of these variants to assess sequencing reads and phenotypic fit highlighted 19 candidate diagnoses (**Table 2**). Of these 19 deletions, all truncated the 5’ UTR and/or immediate upstream promoter sequence and would be predicted to abolish transcription, where the proband phenotype was consistent with homozygous loss-of-function. Three of these ‘likely diagnostic’ non-coding deletions were recurrent: promoter/5’UTR deletions in *EYS, FHL1* and *SIL1* were each seen across two unrelated individuals with consistent phenotype.

**Table 2:**
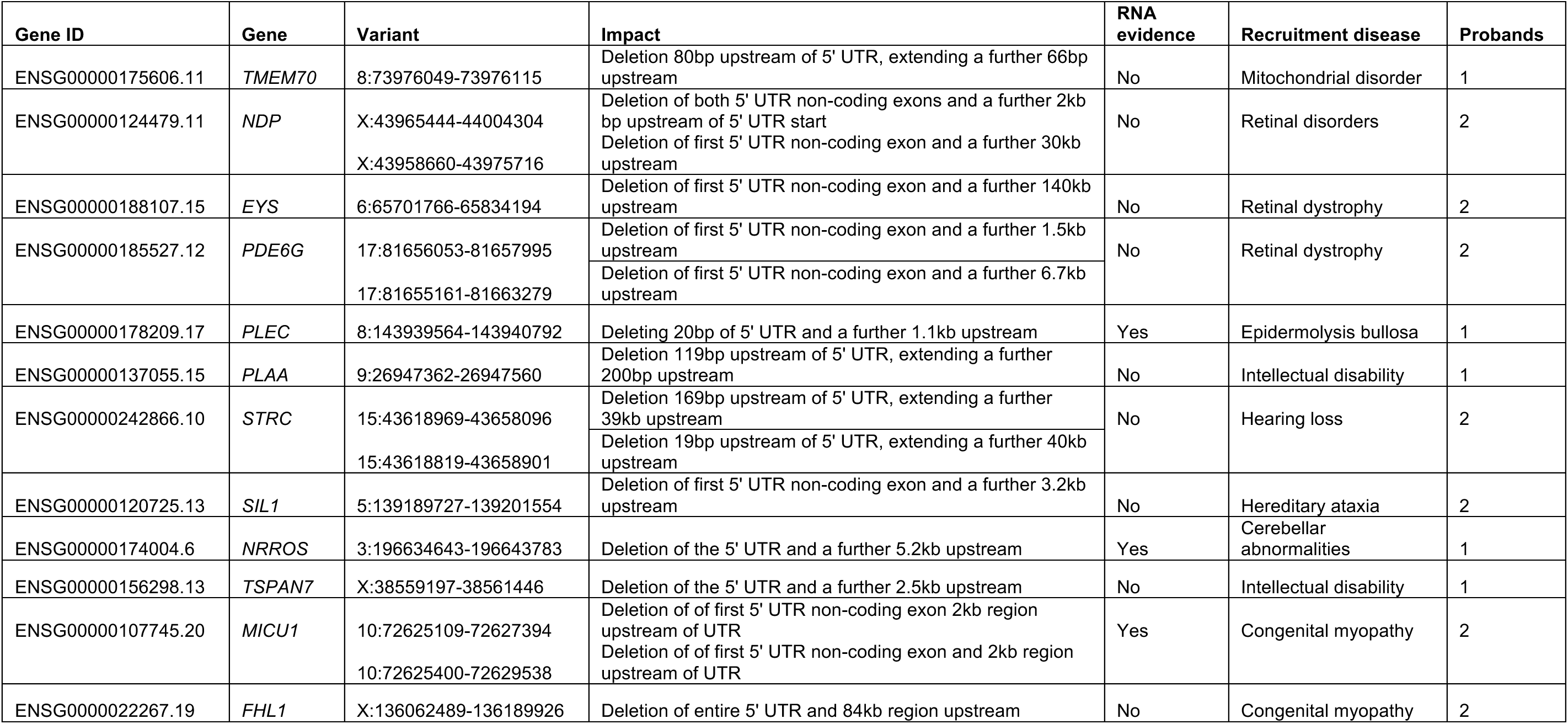
Putative non-coding diagnoses. RNA evidence for *NRROS* was not available in the NGRL for us to review personally, but this case was reported as solved with confirmation of the SV as pathogenic following RNA-sequencing by a diagnostic laboratory. Only the individual with the *NRROS* deletion was marked as solved in the NGRL.

| Gene ID | Gene | Variant | Impact | RNA evidence | Recruitment disease | Probands |
| --- | --- | --- | --- | --- | --- | --- |
| ENSG00000175606.11 | *TMEM70* | 8:73976049-73976115 | Deletion 80bp upstream of 5' UTR, extending a further 66bp upstream | No | Mitochondrial disorder | 1 |
| ENSG00000124479.11 | *NDP* | X:43965444-44004304<br>X:43958660-43975716 | Deletion of both 5' UTR non-coding exons and a further 2kb bp upstream of 5' UTR start<br>Deletion of first 5' UTR non-coding exon and a further 30kb upstream | No | Retinal disorders | 2 |
| ENSG00000188107.15 | *EYS* | 6:65701766-65834194 | Deletion of first 5' UTR non-coding exon and a further 140kb upstream | No | Retinal dystrophy | 2 |
| ENSG00000185527.12 | *PDE6G* | 17:81656053-81657995<br>17:81655161-81663279 | Deletion of first 5' UTR non-coding exon and a further 1.5kb upstream<br>Deletion of first 5' UTR non-coding exon and a further 6.7kb upstream | No | Retinal dystrophy | 2 |
| ENSG00000178209.17 | *PLEC* | 8:143939564-143940792 | Deleting 20bp of 5' UTR and a further 1.1kb upstream | Yes | Epidermolysis bullosa | 1 |
| ENSG00000137055.15 | *PLAA* | 9:26947362-26947560 | Deletion 119bp upstream of 5' UTR, extending a further 200bp upstream | No | Intellectual disability | 1 |
| ENSG00000242866.10 | *STRC* | 15:43618969-43658096<br>15:43618819-43658901 | Deletion 169bp upstream of 5' UTR, extending a further 39kb upstream<br>Deletion 19bp upstream of 5' UTR, extending a further 40kb upstream | No | Hearing loss | 2 |
| ENSG00000120725.13 | *SIL1* | 5:139189727-139201554 | Deletion of first 5' UTR non-coding exon and a further 3.2kb upstream | No | Hereditary ataxia | 2 |
| ENSG00000174004.6 | *NRROS* | 3:196634643-196643783 | Deletion of the 5' UTR and a further 5.2kb upstream | Yes | Cerebellar abnormalities | 1 |
| ENSG00000156298.13 | *TSPAN7* | X:38559197-38561446 | Deletion of the 5' UTR and a further 2.5kb upstream | No | Intellectual disability | 1 |
| ENSG00000107745.20 | *MICU1* | 10:72625109-72627394<br>10:72625400-72629538 | Deletion of first 5' UTR non-coding exon 2kb region upstream of UTR<br>Deletion of first 5' UTR non-coding exon and 2kb region upstream of UTR | Yes | Congenital myopathy | 2 |
| ENSG0000022267.19 | *FHL1* | X:136062489-136189926 | Deletion of entire 5' UTR and 84kb region upstream | No | Congenital myopathy | 2 |

For two probands (with deletions involving *MICU1* and *PLEC*), RNA-sequencing data were available in the NGRL enabling us to study the impact of the variant, with a further proband (with an *NRROS* deletion) already having had the SV confirmed as pathogenic following RNA-sequencing by a diagnostic laboratory.

We identified two unrelated probands with 5’UTR and promoter deletion in *MICU1*. One of these probands had available RNA-sequencing data, which showed a dramatic reduction in *MICU1* transcript levels (TPM) compared to the rest of the cohort (n=7,829, two-sided Z-test; p=7.7x10-5, Z=-3.95 (**Fig. 2C**)), consistent with abolition of transcription. Biallelic loss-of-function of *MICU1* is known to cause myopathy and NDD. The proband exhibited features consistent with *MICU1* deficiency (HPO terms “Hypotonia” and “Congenital myopathy”) supporting this deletion as the correct molecular diagnosis^43^.

Another proband had a promoter deletion of the MANE Select transcript in *PLEC* (ENST00000345136.8, NM_201384.3). In this individual, total expression of *PLEC* remains relatively high in blood (61.03 TPM compared to median of 62.21 in the whole cohort). The transcript structure of *PLEC* is, however, complex, with at least eight alternative first exons, which can be selectively spliced onto a shared set of common downstream exons^44^. Many of these variable first exons lie close to CpG islands, supporting the existence of multiple promoter regions regulating *PLEC* expression^44^.

The most highly expressed *PLEC* transcript in GTEx^45^ does not include the first exon of the MANE Select transcript. RNA-sequencing in the proband does show aberrant splicing, with a reduction in expression of exon 1 of the MANE Select transcript (ENSE00001373903) and loss of the splice junction between exons 1 and 3 (ENSE00001373903 and ENSE00003693086; **Fig. 2D**). Comparing per exon expression revealed that this proband had a depletion of exon 1, with the ratio of exon 1 TPM compared to the mean TPM across all other *PLEC* exons being the 6^th^ lowest of all probands with RNA-sequencing data (7824th of 7829). There is a single pathogenic variant in ClinVar in exon 1 (NM_201384.3:c.46C>T, p.Arg16Ter, VCV000204402.41) which was first reported in two Turkish sisters^46^. The phenotype in these two sisters of epidermolysis bullosa simplex with nail dystrophy is consistent with the phenotype in the proband with the homozygous deletion. Also consistent is the absence of muscular dystrophy and cardiomyopathy which are commonly observed in individuals with loss-of-function variants in exons impacting all plectin isoforms.

We then expanded our search to all non-coding regions (i.e., not restricted to UTRs and upstream promoter regions), looking for overlapping deletions in at least two probands with related phenotypes. However, we did not identify any non-coding biallelic deletions matching these criteria. Combining the 19 non-coding 5’UTR/promoter deletions with the protein-coding deletions described above bring the total diagnostic rate due to biallelic deletion SVs to 0.5% of all rare disease affected individuals in the NGRL (**Supplemental Fig. 7**).

### Biallelic deletions identify novel disease genes

We next sought to leverage our large cohort to discover as-yet unknown gene-disease relationships. We identified 39,444 genes without any OMIM or PanelApp disease annotation, of which 16,078 were protein-coding and 23,366 were non-coding genes. Across these 39,444 genes, 33,109 did not have any exonic biallelic deletions in control individuals (cancer germline or unaffected participants), forming our search-space for novel biallelic disease genes (see **Methods**). We identified 1,572 biallelic deletions in 1,417 individuals that were predicted to disrupt 1,709 of the 33,109 genes (502 protein-coding and 1,198 non-coding genes), without the SV also overlapping any OMIM/PanelApp disease genes. Of the 1,709 genes, 1,391 (81.39%) were only impacted in a single individual. After review, 43 genes harbored biallelic deletions in at least two individuals (72 total participants) with a similar phenotype, which included a consistent recruited disease, HPO term or rare disease relevant ICD-10 code (**Supplemental Table 4**).

To increase our power for gene-discovery, we expanded our analysis to consider small homozygous and compound heterozygous variants with predicted loss-of-function (pLoF) effects (i.e., nonsense, frameshift, and essential splice site variants) in any of the 502 OMIM/PanelApp unannotated protein-coding genes that had at least one biallelic deletion. This approach reidentified the recently published retinal disease gene *SLC66A1*^47^, where five probands with putative pathogenic homozygous variants; two had deletions (2/5) while three had pLoF homozygous SNV/Indels affecting *SLC66A1* across the NGRL.

We also identified a further two genes, *ARHGAP32* and *ARB2A,* where there is one proband with a homozygous deletion and another with a homozygous pLoF SNV. For both genes, each proband has an intellectual disability phenotype. Both *ARHGAP32* and *ARB2A* are expressed in the brain and homozygous pLoF SNVs are absent from UK Biobank^48^ (UKB) and gnomAD^49^ v4.1, with the exception of a single splice donor variant in *ARHGAP32* (chr11:129123886 A>C). This is the donor site to an exon with low proportion expressed across transcripts (pext^50^) score, 0.1 (both across all tissues and in brain), suggesting this SNV may not cause *ARHGAP32* loss-of-function.

We additionally shared the list of genes with at least one homozygous deletion with collaborators with access to large rare disease cohorts (see **Methods**). This highlighted additional cases with *GCG* and *ENTPD3* homozygous deletions or pLoF variants with consistent patient phenotypes (**Supplemental Table 3**).

Three of the 43 genes with homozygous deletions and/or pLoF SNVs in multiple unrelated individuals are discussed in the following sections.

### PDC as a novel gene for Leber Congenital Amaurosis

Four probands from four families in the NGRL cohort are homozygous for a 15kb deletion that contains the entire canonical transcript of *PDC* (ENST00000391997; **Fig. 3A**). The deletion was confirmed in one family by RT-PCR (**Supplemental Fig. 8**) and was identified as heterozygous in an unaffected sibling in one family (**Supplemental Fig. 9**). The four affected individuals with the homozygous deletion have early-onset, rapidly progressive retinal degeneration: one patient was diagnosed with Leber congenital amaurosis in infancy and had lost all light perception in their 20’s (**Supplemental Fig. 10**). There were a further 13 heterozygote carriers of this allele in the NGRL with no recorded eye manifestations, supporting a recessive loss-of-function disease model. *PDC* encodes phosducin, which plays a key role in the visual phototransduction cascade by binding the β and γ subunits of transducin, a G-protein that links activated rhodopsin to cGMP-phosphodiesterase during light detection^51^ (**Fig. 3B**). Through modulation of transducin subunit availability, phosducin contributes to the regulation of phototransduction signaling. Transcriptomic analysis of 201 post-mortem eye samples^52^ confirmed that the canonical transcript of *PDC* is highly expressed in the adult human retina (mean TPM = 351, SD = 179) and retinal pigment epithelium (mean TPM = 258, SD = 132) (**Supplemental Fig. 11**).

**Figure 3:**
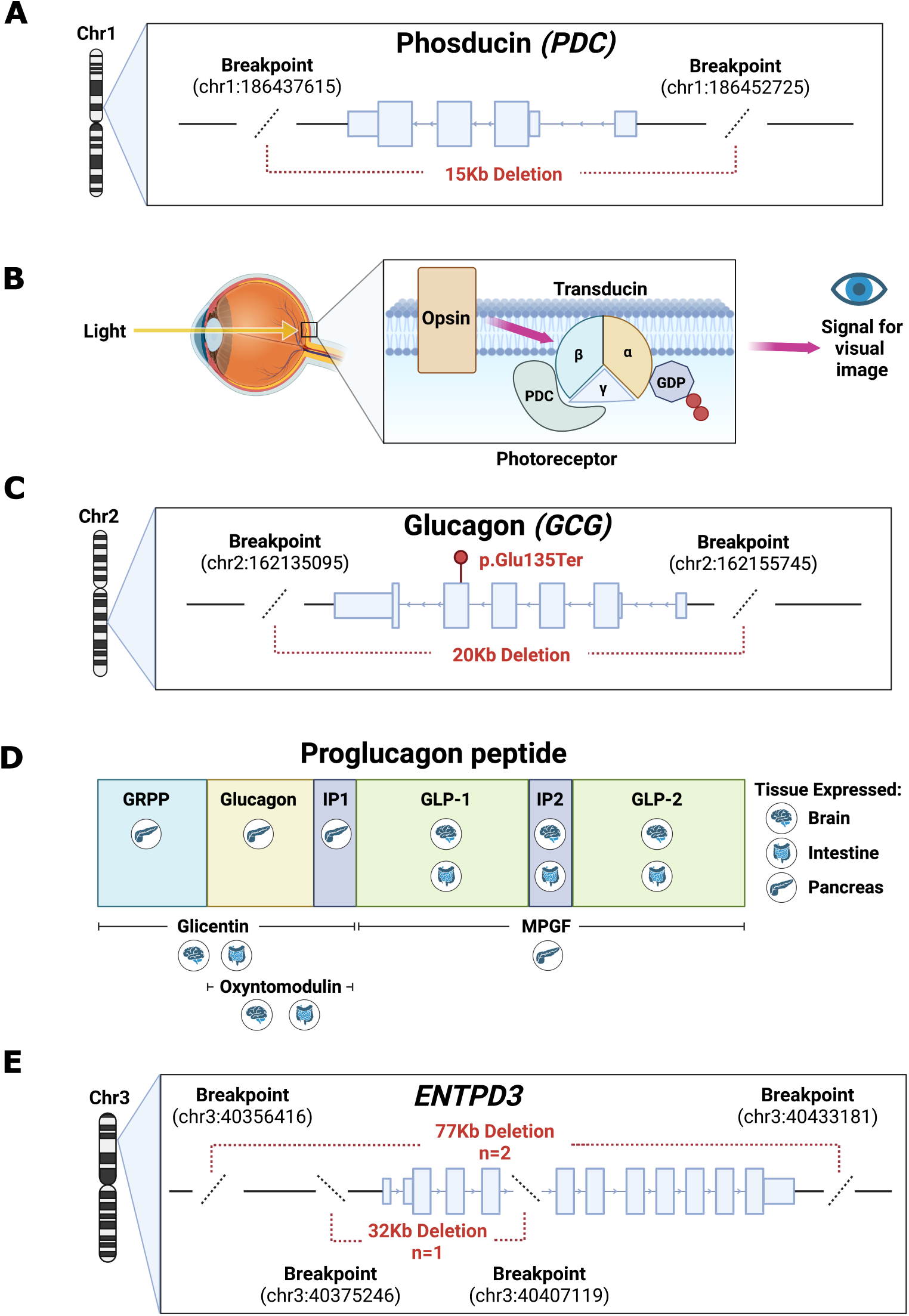
*PDC* and *GCG* are novel gene-disease relationships with recessive inheritance. **(A)** Schematic of the 15kb *PDC* deletion seen in four families in the NGRL which encompasses the entire gene. (**B)** Diagram showing *PDC*’s role in visual phototransduction. It binds to the β and γ subunits of the transducin complex. **(C)** Schematic showing putative pathogenic variants in the two sisters from the NGRL (homozygous 20Kb deletion) and a proband from an Australian rare disease cohort (homozygous stop-gain SNV, ENST00000418842.7:p.Glu135Ter) with a similar phenotype. **(D)** The nine different peptides produced by preproglucagon mRNA cleavage in a tissue specific manner are illustrated alongside the respective tissue(s) that peptide is expressed in. **(E)** Schematic of the two *ENTPD3* deletions seen across the 3 probands. The 32Kb and one of the two 77Kb homozygotes are from the NGRL cohort, whilst the second 77Kb deletion proband coming from the NHGRI GREGoR cohort (see **Methods**).

In the UK Biobank cohort^53^ we identified 307 individuals with the *PDC* deletion in the heterozygous state (allele frequency (AF) of 1 in 3,096), alongside 5 in gnomAD SV (AF of 1 in 25,218). We investigated SNVs in-cis with the deletion allele to construct a *PDC* haplotype. A nearby SNV (chr1:186,724,165 A>T; GRCh38) was present in 310 UKB heterozygotes, of whom 307 also had the *PDC* deletion on the same allele. Comparing frequency of this SNV between the UKB and gnomAD non-Finnish Europeans, there is significant enrichment in UKB, suggesting that the *PDC* deletion could be a UK founder variant (UKB 307/980,773, gnomAD v4.1 non-Finnish EUR 7/68,019; Chi-Square test, OR = 3.04; *P* = 3.20×10^-3^). Mapping the UKB carriers’ place of birth in the UK suggests the variant may have originated in north-west England (**Supplemental Fig. 12**) roughly 38 generations (600-1000 years) ago^54^.

### GCG is a novel gene for intellectual disability with gastrointestinal involvement

We identified two sisters with a homozygous deletion encompassing the entire *GCG* gene (chr2:162,135,095-162,155,745; **Fig. 3C**). Their symptoms include intellectual disability, recurrent infantile diarrhea, and gastrointestinal issues. Both unaffected parents have the deletion in the heterozygous state.

While we did not identify any other individuals in the NGRL cohort with biallelic deletions or SNVs impacting *GCG*, we uncovered homozygous stop gain variant (NM_002054.5:c.403G>T; p.Glu135Ter) in a proband through the RDNow cohort (see **Methods**). This individual presented with autism, delayed speech and language development, along with infantile-onset diarrhoea that resolved in later years. Hence, all three individuals have predicted biallelic loss of *GCG* and a phenotype involving intellectual disability and gastrointestinal issues including infantile-onset diarrhea.

*GCG* encodes proglucagon, a preprohormone peptide that is subsequently cleaved in a tissue specific manner to produce multiple smaller functional peptides: glucagon, GLP-1 and GLP-2^55,56^ (**Fig. 4D**). The importance of these peptides in both the brain and intestine aligns with the tissue-specific phenotypes observed in these two individuals.

### ENTPD3 as a novel gene for intellectual disability and autism

We identified two probands in GEL with distinct homozygous deletions in *ENTPD3*. The smaller SV (chr3:40375246-40407119) removed 32Kb of sequence including the first four exons of the MANE Select transcript (ENST00000301825.8, NM_001248.4). The larger SV (chr3:40355945-40433833) deletes the entirety of *ENTPD3* (**Fig. 4E**). Neither SV is present as homozygous in gnomAD v4.1 (AF of 7.1x10^-5^ and 1.1x10^-4^ for the small and large deletions respectively). There is only a single individual in gnomAD v4.1 with a homozygous pLoF variant in *ENTPD3* (splice donor variant chr3:40411964 T>G), however, this individual has lower coverage at this position than most heterozygous carriers, with one of the 16 reads not containing the variant. Both probands were recruited under ‘intellectual disability’ and shared a consistent phenotype; intellectual disability, microcephaly (HPO ‘microcephaly’) and autism (HPO ‘Autistic Behaviour’; ICD-10 ‘Childhood Autism’). We uncovered a third proband from the NHGRI GREGoR cohort (see **Methods**) with the larger deletion with a consistent phenotype of intellectual disability and autism. *ENTPD3* is highly expressed in the brain: in GTEx four of the five tissues with highest *ENTPD3* expression are brain tissues (e.g. the caudate (basal ganglia) with a TPM of 17.36). Collectively, these data are consistent with homozygous loss of *ENTPD3* as a novel cause of intellectual disability.

## Discussion

Structural variants are a major driver of rare disease, yet their identification and annotation remain challenging with short-read sequencing data. By enriching for the read-depth features characteristic of homozygous deletions, we generated a genome-wide high-confidence dataset of homozygous deletions across 125,730 individuals.

These data enabled us to investigate genome-wide resilience to homozygous loss, highlighting that a substantial proportion of the genome, including 0.18% of protein-coding sequence, is tolerant to homozygous knockout. Such a map of biallelic deletions has broad utility outside of clinical diagnosis in rare disease. For example, understanding which genomic regions are tolerant to complete loss is insightful for therapeutic approaches which aim to decrease a gene product. In such cases this provides evidence that knocking down the gene product may not be deleterious, adding to similar evidence from studies focusing on small pLoF variants^57^.

A substantial proportion of disease burden is driven by homozygous deletions, amounting to 0.5% of the NGRL cohort. This fraction may also be an underestimate of the overall disease burden from this category of variants as individuals were often only recruited to the NGRL after prior negative testing using assays (e.g., microarrays) that may identify deletion-based diagnoses. Indeed, a third (113/322, 35.09%) of the likely-diagnostic deletions identified were smaller than the size-limit for detection via ultra high-resolution microarray (∼10Kb)^58,59^, which increases to almost half (49.07%) when using a more typical limit of 30Kb. The candidate diagnoses that we identified included SVs in known recessive disease genes that only impact non-coding regions. Such SVs are not routinely annotated in clinical settings or may be discarded in bioinformatics pipelines given that they do not impact protein-coding regions. In particular, we find multiple examples of deletions of the start of the 5’UTR and the upstream promoter that would be predicted to cause complete loss of gene transcription. Similar deletions have been identified in haploinsufficient genes, including including *MEF2C*^60^ and *MYBPC3*^61^, and our data support this as an underappreciated pathogenic mechanism.

We also demonstrated the utility of homozygous deletions to discover novel recessive disease genes. Although large deletions often span multiple genes, hindering precise gene attribution and hence novel gene discovery, we highlight three cases where a single deletion completely eliminates just one gene. One example is in *PDC*, which we propose as a novel disease-association with Leber congenital amaurosis. Given its role within the phototransduction cascade, *PDC* has long been considered a strong candidate gene for inherited retinal disease; however, prior mutation screening studies in retinal disease cohorts have not identified convincing pathogenic variants, and a definitive human phenotype had not been established until now^62^.

We show that this PDC deletion could be a UK founder variant, which likely accounts for additional undiagnosed cases in the broader population. Phosducin’s compact coding sequence easily fits within the ∼4.7 kb packaging limit of adeno-associated virus (AAV) vectors. Notably, AAV-based gene therapy for Leber congenital amaurosis (targeting RPE65) has been FDA-approved since 2008, providing a precedent for this approach.

The youngest of our PDC probands was evaluated for AAV-mediated gene replacement therapy but unfortunately retinal degeneration has already progressed beyond the point where preventing further damage could be clinically beneficial.

We also highlight *GCG* as a candidate novel gene-disease association. *GCG* encodes several key peptides, including its namesake glucagon, yet surprisingly complete loss of these hormones is compatible with life. Although the precise contribution of each peptide to the observed symptoms remains unclear, the gastrointestinal dysfunction in the two affected sisters strongly implicates GLP-1 deficiency. From a therapeutic standpoint, administering exogenous GLP-1 or related analogs may markedly improve their GI symptoms. To validate this finding and to understand each hormone’s role in *GCG* loss-of-function, we are now conducting a *GCG* knockout mouse study coupled with targeted hormone replacement. Finally, we identified three probands with intellectual disability with two different homozygous deletions in *ENTPD3*. While the near absence of homozygous pLoF variants in gnomAD and high expression of *ENTPD3* in the brain are supportive, replication in additional cohorts is needed to confirm this as a novel recessive ID gene.

Our study has several limitations. Despite our careful approach to identify and annotate SVs, our dataset likely still contains false positive SVs. Indeed, during our manual curation of SVs impacting known disease genes, around 0.1% were determined as unlikely true SV calls, or to be unlikely to actually truncate the coding sequence of a gene, when visualising sequencing reads on IGV. Conversely, our strict read-depth filter may have removed true positive SV calls, although notably we did not miss any known diagnostic biallelic deletions in the NGRL cohort. We are also limited by our use of short-read data and may miss larger SV events that will need long-read sequencing to accurately detect. We describe three genes with biallelic deletions in at least three individuals as putative novel disease genes, however, there are likely to be other diagnoses within our list of 43 genes with one or two individuals with biallelic deletions. This is highlighted by the abundance of n of 1 diagnoses even in known recessive disease genes in the NGRL. Finally, it remains difficult to annotate variants in non-coding regions. Here, we focused on recurrent variants and those overlapping known disease genes, given that they are far easier to interpret. Our future work will investigate singleton variants in intergenic regions that may impact cis-regulatory elements and/or genome architecture.

Further, our list of genes without a known disease association but with deletions in one or more NGRL individuals will be useful for analysis in other cohorts, increasing power to discover additional novel recessive gene disease associations. In summary, we have generated a high-confidence dataset of homozygous deletions. These data give insights into the genome-wide burden and tolerance of homozygous deletion, underscore the significant role of coding and non-coding biallelic deletions in human disease, and enable discovery of novel disease genes.

## Data Availability

Data from the National Genomic Research Library (NGRL) used in this research are available within the secure Genomics England Research Environment. Access to NGRL data is restricted to adhere to consent requirements and protect participant privacy. Data used in this research include:
Structural variant vcfs for SV calls from the NHS-GMS and 100,000 Genomes Project participants, and their BAM files for coverage calculation, with file paths available in the genome files and paths labkey table.
Phenotype data, including HPO terms, ICD-10 codes and recruited disease information, were taken from the hospital episode statistics and participant phenotype labkey tables.
AggV3 aggregate SNV data was used to discover SNV/Indel predicted loss-of-function variants as orthogonal evidence for putative novel disease genes.
Access to NGRL data is provided to approved researchers who are members of the Genomics England Research Network, subject to institutional access agreements and research project approval under participant-led governance. For more information on data access, visit: https://www.genomicsengland.co.uk/research

## Acknowledgements

We gratefully acknowledge the participants of the National Genomic Research Library (NGRL), whose contributions made this research possible. Secure access to the NGRL under project ID RR566 was provided by Genomics England, which delivers the NGRL in partnership with NHS England, and is wholly owned by the UK Department of Health and Social Care. The NGRL contains participants’ health data collected by the NHS as part of their care, along with samples and data from their participation in research, for which fully informed consent has been obtained. This includes genomic and clinical data provided through the NHS Genomic Medicine Service, as well as data obtained through research studies, including the 100,000 Genomes Project and the Generation Study, both of which are delivered in partnership with the NHS, and from other research cohorts involving external collaborators.

This research has also been conducted using the UK Biobank Resource under Application Number 103356 and uses data provided by patients and collected by the NHS as part of their care and support.

A.M. is supported by a Wellcome Trust Four-year PhD Studentship in Basic Science. N.W. is supported by a Wellcome Career Development Award (grant no.305292/Z/23/Z) and a Lister Institute research prize. This work was supported by the Medical Research Council Centre of Research Excellence in Therapeutic Genomics (grant number MR/Z504725/1 to S.J.S., J.T., N.W.) and Health Data Research UK QQ2 Molecules to Health Records Driver Programme to S.J.S. A.O.D.L. is supported by National Human Genome Research Institute grants U01HG011755 and R01HG013986. L.E.C. is supported by a Manton Center for Orphan Disease Research postdoctoral fellowship.

Figure 3 graphic was created with the help of BioRender and licensed for use in publication under agreement number IA29OVE394.

## Methods

### Creation of a high-confidence deletion set

Structural variant VCF files were obtained from the latest version of NHS-GMS and 100kGP data available in the NGRL at the time of this analysis; v2024_31_12. The VCF and BAM/CRAM files along with relevant metadata (e.g. participant id) were obtained using Labkey API queries (v1.04). Bcftools^63^ v4.01 was used to obtain homozygous CNV/Deletion calls, retaining variants that failed QC filters inbuilt into the detection tool. Pre-depth filtering breakdowns based on genome build, cohort and variant caller are shown in **Supplemental Table 5**.

Samtools^63^ v1.1 was used to calculate the mean and median read depth across each deletion. The median read depth across 100 random (unrelated) participants was used to generate a control read depth across every putative deletion. Controls were sampled from unaffected family members of rare disease recruited probands and participants recruited for cancer (where there were ‘somatic’ and ‘germline’ .bam files available from joint tumour/normal sequencing, only the ‘germline’ was utilised). For chromosome X, only females were used as controls, as haploid males would artificially lower control coverage compared to the autosomes.

A unified dataset of SV calls was generated across 100kGP (GRCh38/GRCh37) and NHS-GMS (GRCh38). For a variant to be considered it needed (1) A median coverage across the variant of ≥ 10 reads in the controls, and (2) A median coverage of ≤ 1 read in that participant. As such, a variant could be included in one individual and excluded in another depending upon the coverage observed across the variant call in both cases. Variants were grouped based on ≥80% overlap. Allele frequencies for the SVs were calculated post-overlap grouping.

Duplicate participants between the 100,000 Genomes Project and the NHS Genomic Medicine Service were flagged from an internally generated list of duplicate participant IDs (n=847) based on KING relatedness^64^, and removed to prevent double counting.

Out of the 332 likely diagnostic variant calls, 126 (37.95% of the total diagnoses) failed QC filters inbuilt into the detection tool but passed our homozygous deletion specific read-depth based filtering approach. This is despite our approach retaining fewer variants than tool-based PASS filtering, suggesting that our approach enriches for true positive SV variants. 31 variants failed QC solely for low quality, 82 fell below the 10kb minimum deletion size imposed by Canvas and 23 met both exclusion criteria.

All candidate diagnostic or putative novel disease gene variants were reviewed manually in Integrative Genomics Viewer^65^ (IGV) to prevent misattribution of caller or sequencing artefacts as disease relevant SVs.

### Variant annotation and identification of candidate diagnostic SVs

Variants were annotated using (1) VEP^66^ v112 for predicted impact; (2) Labkey for participant phenotype information (i.e. HPO terms and recruited disease) (3) OMIM^36^ and PanelApp^35^ for OMIM ‘morbid’ and PanelApp ‘green’ respectively. PanelApp genes were those with the evidence field as ‘green’ and mode of inheritance containing either the term ‘recessive’ or ‘X-linked’; this returned 2,973 genes. OMIM recessive genes were those with a HPO of ‘recessive inheritance’, ‘x-linked recessive inheritance’, ‘x-linked inheritance’ or ‘‘x-linked inheritance’; this returned 3,239 genes. These were combined into a total set of 3,580 unique disease genes where complete loss-of-function may be expected to cause a rare disease phenotype. The PanelApp and OMIM annotations were taken from the latest versions as of 16^th^ August 2025.

Variants were annotated as potential promoter knockout if they overlapped a 250nt window upstream of the most 5’ UTR based on the MANE^67^ Select transcript for that gene without overlapping its protein-coding sequence, using bedtools^68^ v2.03.

Where a variant was predicted to truncate a disease gene by an SV caller without base-pair resolution (i.e. Dragen and Canvas), bedtools was used to intersect the putative variant location with MANE features of interest (e.g. coding exons). The overlapping region was then rescanned for coverage with a median read-depth of ≤1 being used as the threshold for retention for manual review in IGV.

A variant was taken to be a diagnosis if: (1) it was predicted to be truncating (either whole gene deletion or frameshift) the protein-coding sequence or promoter region of a biallelic disease gene, (2) it was not present in any individuals who were recruited for cancer or unaffected relatives of a rare disease proband 3) The phenotype matched what one would expect for biallelic loss-of-function in the gene, as taken from OMIM and PanelApp. An additional diagnosis was taken to be one where the case was annotated as partially solved within the NGRL and the deletion would explain the clinical features unresolved by the first diagnosis.

### Annotation type of deleted nucleotides

For the data shown in **Supplemental Table 1**, annotation information was taken from GENOCDE v49. Overlapping sequence across a given annotation type, such as overlapping ‘UTR’ regions in multiple transcripts or genes, were merged into single intervals using bedtools *merge*. For ‘intronic’ sequence, ‘exonic’ annotations were removed from the gene region (via bedtools *subtract*). In cases where a nucleotide had multiple annotations, (e.g. coding in one transcript and UTR in another) it was included in the nucleotide count of all relevant categories.

### Novel gene discovery

To define the novel disease gene search space, we complied all unique gene annotations from NCBI’s RefSeq^69^ transcript data as of 16th August 2025. This gave a list of 43,024 unique genes (with 20,078 or 46.7% protein-coding). We then removed any which were present in our OMIM and PanelApp disease gene set as described in the diagnostic SV section above, leaving a total of 39,444 recessive disease gene candidates.

Of these 24,446/39,444 were predicted to overlap exons (coding or non-coding) in the high confidence set. Of the variants with a predicted impact in these 24,446 genes, we further filtered for cases where the deletion was predicted to truncate an exon (either coding or non-coding). Novel disease genes were those where: (1) the variant was overlapping with a MANE or RefSeq exon annotation (2) the variant was not responsible for another molecular diagnosis (i.e. a SV disrupting a known disease gene, which is diagnostic, alongside other genes without a disease annotation) (3) the variant was present in at least two individuals with a similar phenotype, defined as either the recruited disease category, or, a HPO term matching between all individuals with the variant.

Orthogonal evidence of gene constraint (LOEUF^70^), absence of homozygous pLOF’s in population databases (UK Biobank, gnomAD), in which tissue(s) the gene is expressed (i.e. GTEx^45^), and whether there are knockout data available in other organisms^71^ were then considered. The highlighted candidates, *PDC*, *GCG and ENTPD3*, were depleted of homozygous pLOF variants in the population databases and are expressed in tissue congruent with the proband phenotype.

### Identifying SNVs to support novel disease genes

The 100kGP aggregated dataset (Aggv2)^72^ was used to identify homozygous and compound heterozygous pLoF SNVs (annotated with VEP) in the genes within our novel gene ‘search space’ that had at least one individual with a biallelic deletion. The phenotypes of identified individuals were then compared with the SV proband(s), with a pLoF SNV being assumed to have a similar effect to a truncating SV.

### Quantification of gene expression in the human retina

RNA sequencing data of 201 post-mortem eye samples generated by the Manchester Eye Tissue Repository Genome-Transcriptome Project^52^ (available from the European-Genome phenome Archive (EGA; Study ID: EGAS50000001443; Dataset: EGAD50000002082)) were utilised to study expression levels of *PDC* in the adult human retina and retinal pigment epithelium. Transcriptomic data was generated by short read bulk-RNA sequencing of polyadenylated enriched RNA, using an Illumina NovaSeq6000. Alignment of human reference genome GRCh38 was performed using STAR v2.7.4^73^. Gene and transcript-level quantifications in transcripts per million were generated with RSEM v.1.3.0^74^ using GENCODE^75^ v38 gene annotations.

### Investigating the PDC haplotype

The total allele count of the PDC deletion allele was found by using the heterozygous and homozygous SV calls from SVRare^76^, providing an expected allele count for SNVs in-cis with the deletion allele. SNVs within a 200Kb window of either breakpoint which were heterozygous in the *PDC* deletion carriers or homozygous in the probands were taken to be potential haplotype SNVs. The Aggv2 data was used to get the allele counts for these SNVs across the 100,000 Genomes Project cohort. The SNV allele counts of the *PDC* deletion allele cohort were divided by those seen in Aggv2 (wide cohort) to give a ratio for comparison. The closer to 1 this ratio was, the more informative the SNV allele is that the deletion allele is also present. The most informative SNV, 1-186724165-A-T, was validated in an orthogonal dataset, UK Biobank^53^, where it was seen in 307/311 heterozygotes subsequently confirmed to carry the *PDC* deletion allele. This SNV was used as the estimator for the deletion allele in gnomAD, with a Chi-square test being used to compare this SNV allele count in UKB (307/981,080) versus gnomAD non-Finnish Europeans (7/68,026) to demonstrate UK enrichment.

### Additional Cohorts

#### GREGoR

Additional screening was performed in 4668 probands and family members sequenced by the Broad Institute Center for Mendelian Genomics as part of the Genomics Research to Elucidate the Genetics of Rare diseases (GREGoR) consortium^77^, funded by the NHGRI. Participants were recruited through the Rare Genomes Project (RGP) or through external partners.

#### CaRDinal

The Centre for Population Genomics (CPG) CaRDinal cohort is an Australian rare disease metacohort comprising more than 10,000 participants from over 6,000 families enrolled through 40 research projects. The metacohort includes more than 8,000 genomes and over 3,500 exomes. The family relating to the *GCG* variant (NM_002054.5:c.403G>T; p.Glu135Ter) was consented and enrolled under the Rare Diseases Now (RDNow): Genomic Diagnoses and Personalised Care for Children with Undiagnosed Rare Diseases program.

## Data Availability

Data from the National Genomic Research Library (NGRL) used in this research are available within the secure Genomics England Research Environment. Access to NGRL data is restricted to adhere to consent requirements and protect participant privacy. Data used in this research include:

- Structural variant vcfs for SV calls from the NHS-GMS and 100,000 Genomes Project participants, and their BAM files for coverage calculation, with file paths available in the ‘genome files and paths’ labkey table.
- Phenotype data, including HPO terms, ICD-10 codes and recruited disease information, were taken from the ‘hospital episode statistics’ and ‘participant phenotype’ labkey tables.
- AggV3 aggregate SNV data was used to discover SNV/Indel predicted loss-of-function variants as orthogonal evidence for putative novel disease genes.

Access to NGRL data is provided to approved researchers who are members of the Genomics England Research Network, subject to institutional access agreements and research project approval under participant-led governance. For more information on data access, visit: https://www.genomicsengland.co.uk/research

## Code availability

A Nextflow^78^ pipeline (nf_v11.02) was written to undertake the analysis and is available on GitHub (https://github.com/Computational-Rare-Disease-Genomics-WHG/biallelic_sv).

