## Supplemental Figures for "A genome-wide deletion map in 125,730 individuals for novel rare disease gene and variant discovery"

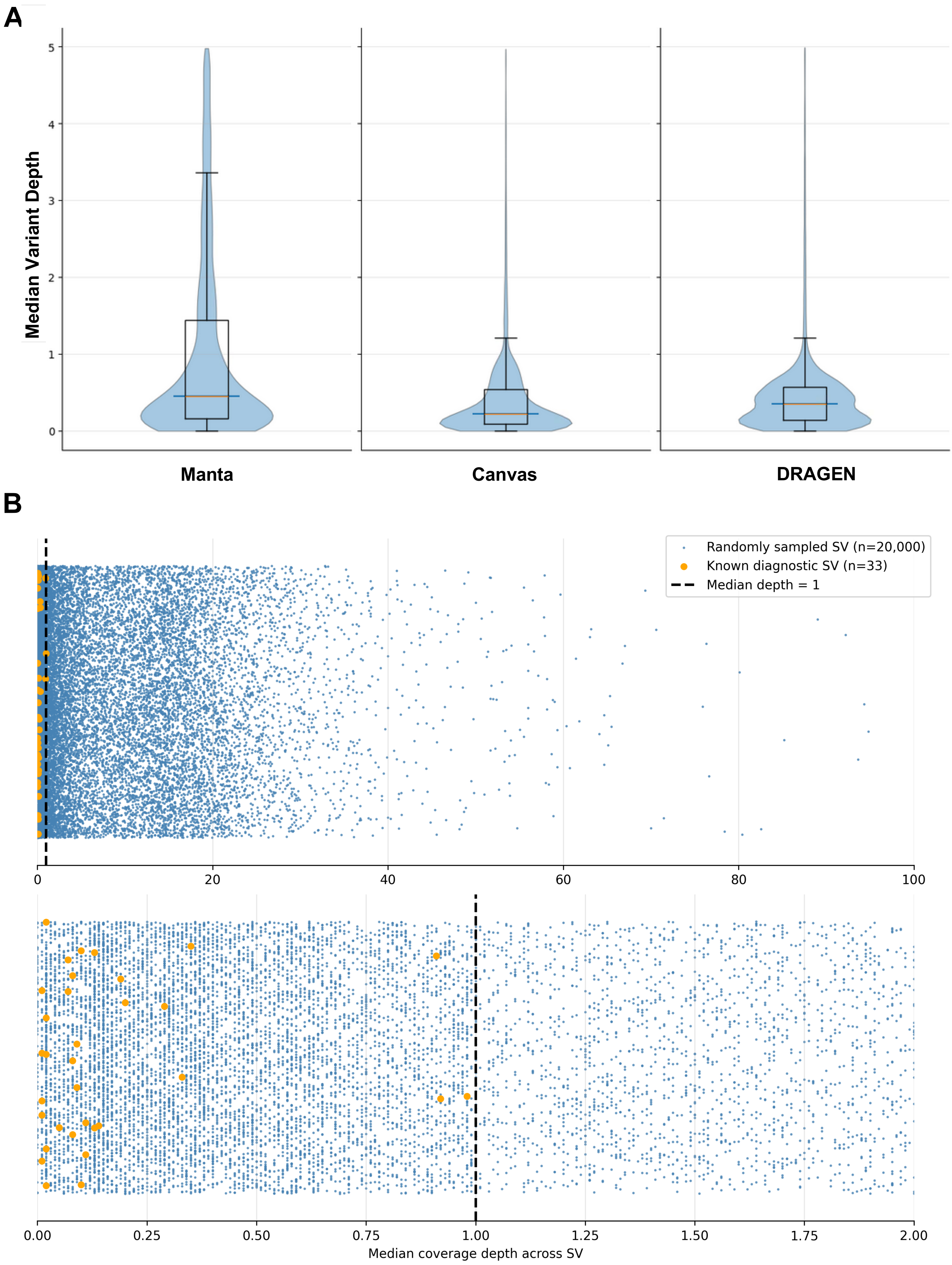


**Supplementary Figure 1: Median depth cutoff determination. (A)** Violin plot of showing the distribution of median coverage depth of putative biallelic deletions across random samples of SVs from each caller (n=20,000). **(B)** Scatter plot median read depth, comparing a random sample (n=20,000) of SVs to diagnostic Rare Disease variants where biallelic deletion is the confirmed molecular diagnosis (n=33).

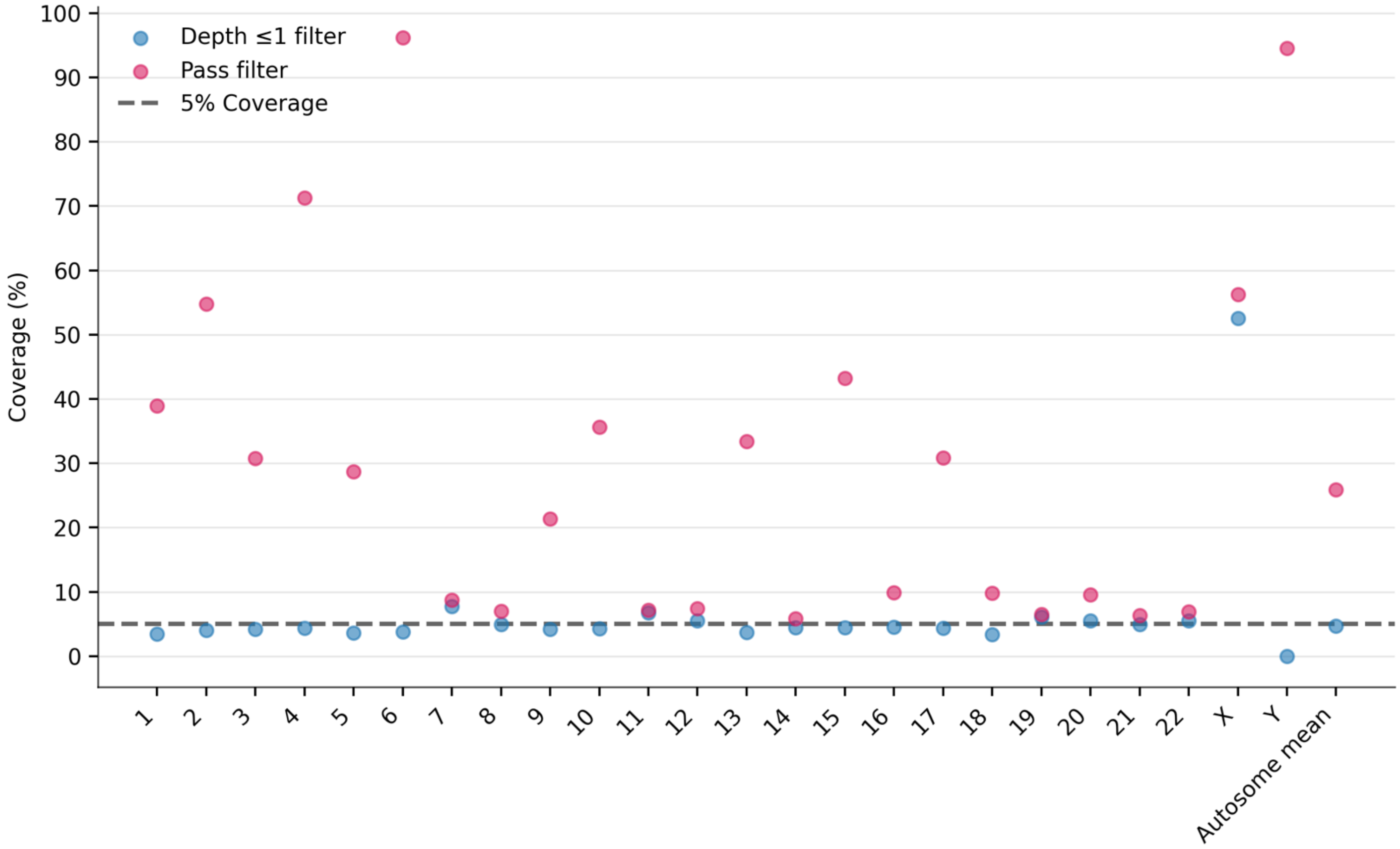


**Supplementary Figure 2: Proportion of unique sequence overlapping a biallelic deletion.** Scatterplot of the unique nucleotide sequence covered by putative deletions, split based on filtering criteria. Median <= 1 filter removes variants when median depth across the putative SV was greater than 1. PASS filter removes SVs that failed any of the respective Manta/Canvas/DRAGEN standard filtering criteria, such as genotype quality >= 15.

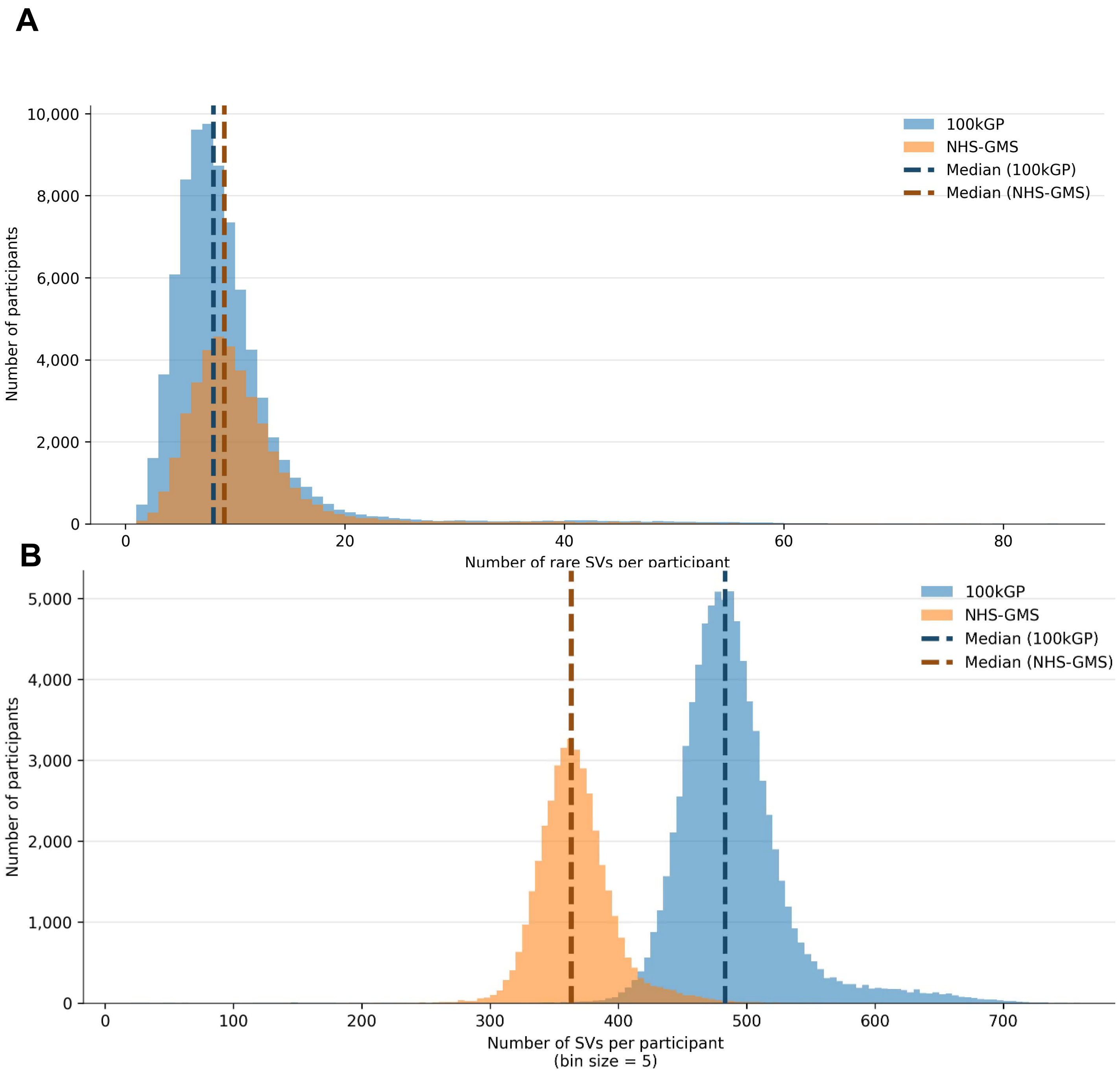


**Supplementary Figure 3: SV distribution by NGRL cohort.** Histograms showing the distribution of SVs for the 100kGP and NHS-GMS cohorts, with median values highlighted. **(A)** All variants, with a bin size of 5. **(B)** SVs filtered for AF ≤ 0.001, with a bin size of 1.

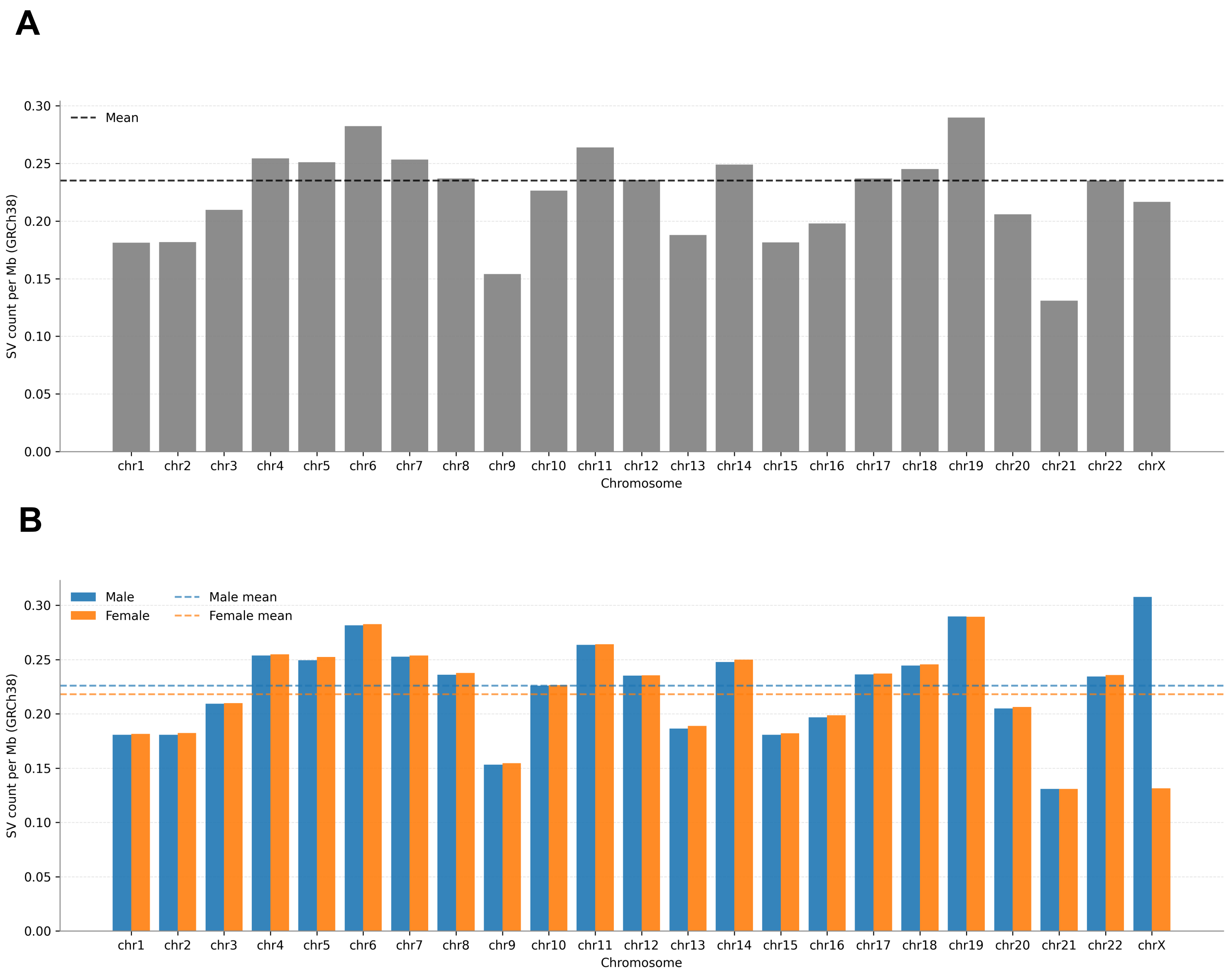


**Supplementary Figure 4: Normalised SV burden per chromosome. (A)** Total high confidence biallelic deletion SV counts per chromosome, normalised based on chromosome size (GRCh38). **(B)** Breakdown of (A) by participant self-reported sex.

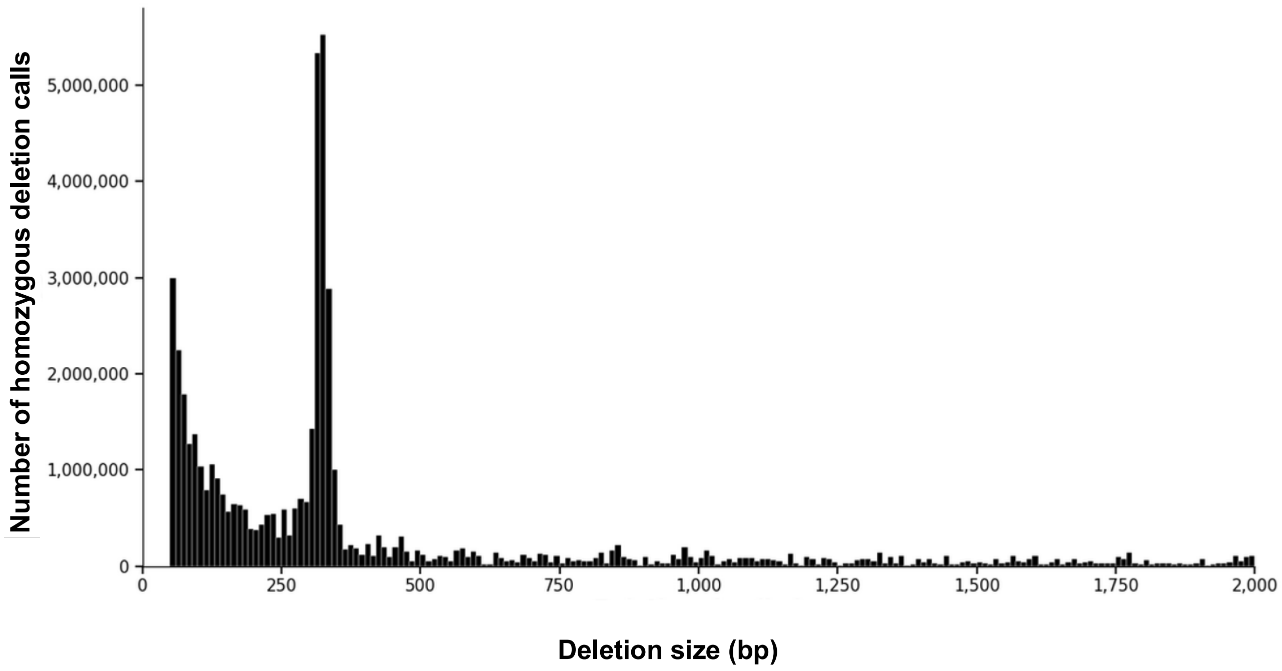


**Supplementary Figure 5: Size distribution of biallelic deletion calls.** Variant frequency histogram based on deletion length, with 10bp window bin size. Includes only variants <= 2Kb in size (n=50,826,503). Deletions >2kb in size (n=13,758,711) are not shown.

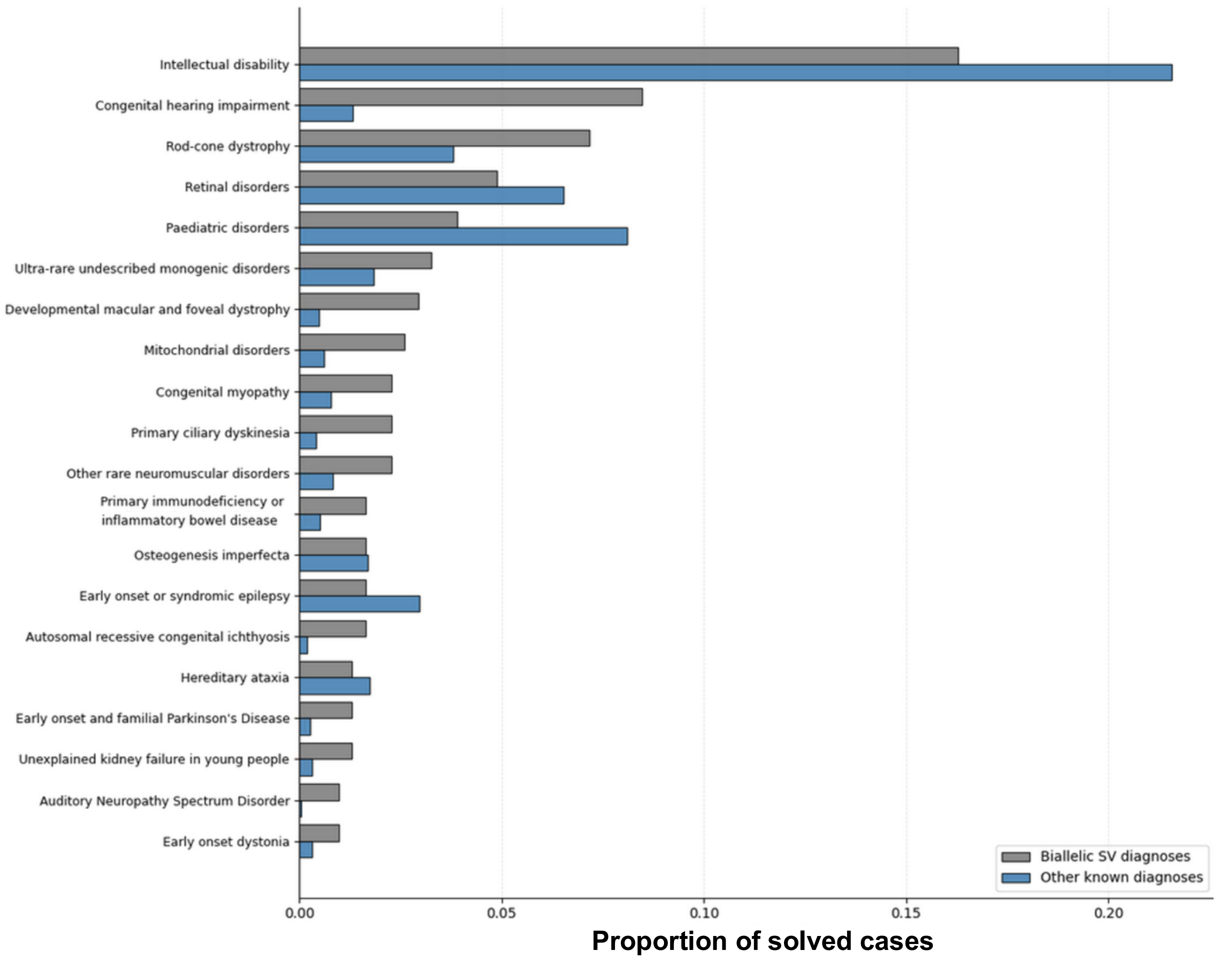


**Supplementary Figure 6: Top 20 diagnoses by recruited disease.** Bar chart showing the proportion of individual recruited diseases with molecular diagnoses in our biallelic SV set, alongside the NGRL cohort as a whole (excluding the biallelic SV putative diagnoses).

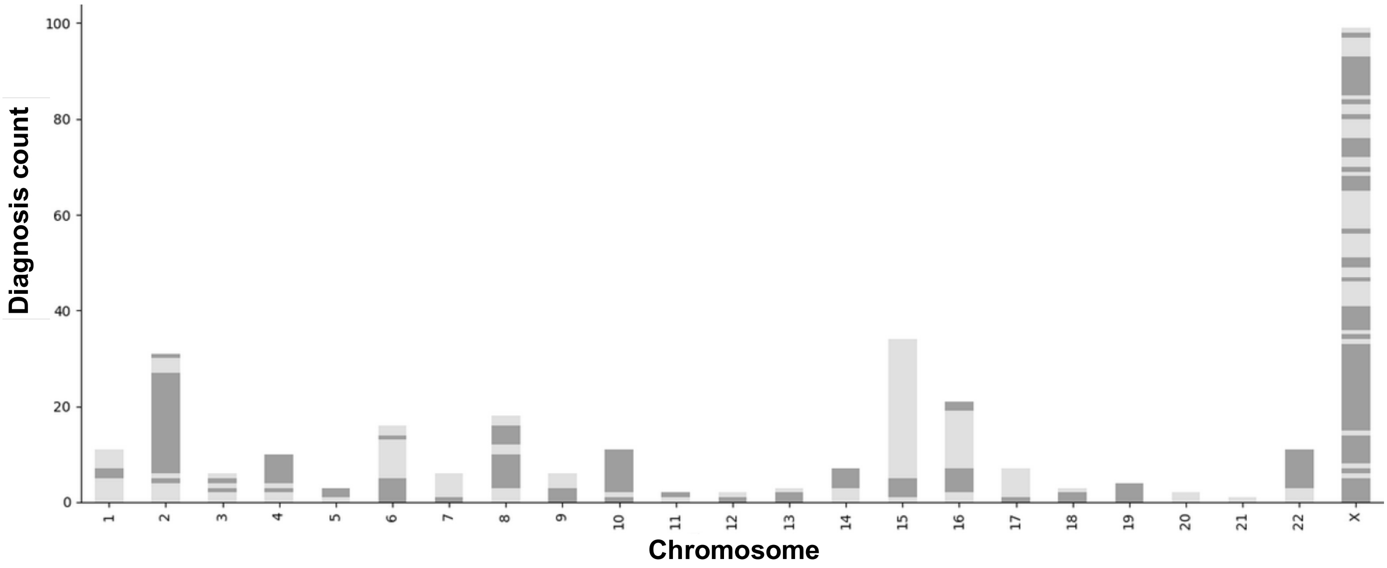


**Supplementary Figure 7: Unique diagnoses per chromosome and gene.** Stacked bar chart showing the number of biallelic deletion related putative diagnoses. Bars are stacked in alternating shades to distinguish different disease-associated genes**.**

**Reference allele**

| Primer pair | Size between primers | PCR band expected |
|---|---|---|
| Longer-F & R | 15,807 bp | None |
| F & R | 15,492 bp | None |
| Internal-F & R | 268 bp | 268 bp band |

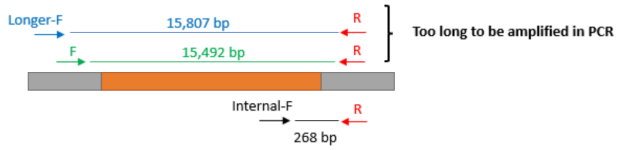


**Allele with DEL**

| Primer pair | Size between primers | PCR band expected |
|---|---|---|
| Longer-F & R | 697 bp | 697 bp band |
| F & R | 382 bp | 382 bp band |
| Internal-F & R | Priming not possible for Internal-F | None |

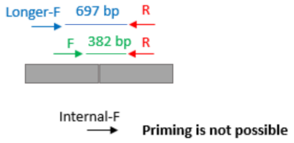


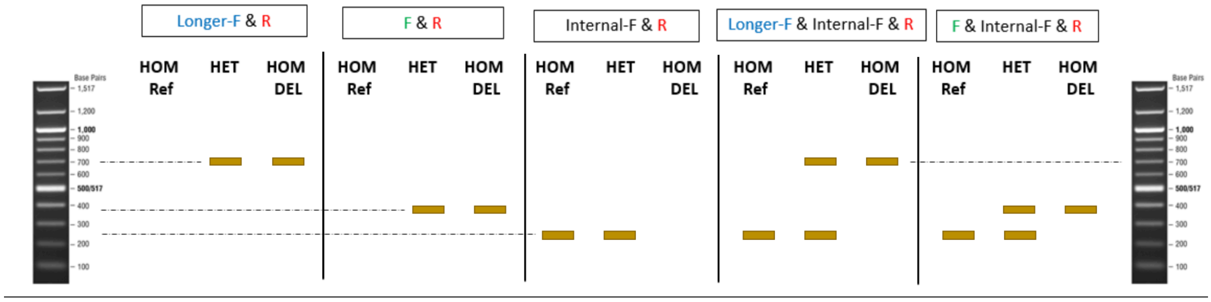


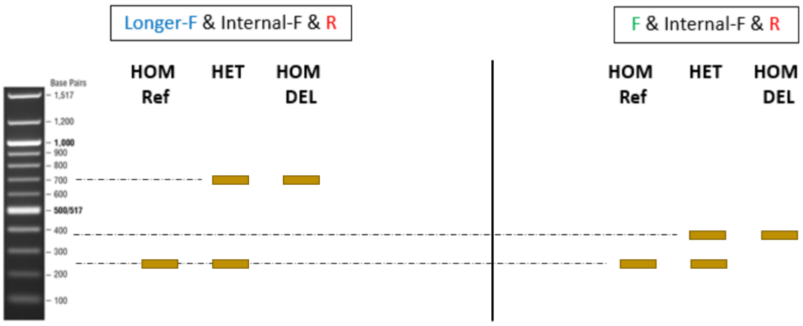


**Samples**

| | |
|---|---|
| CT | Control DNA |
| US | DNA received as AA39236 (unaffected sister of GEL 115009817) |
| 5920 | DNA received as 1010045920 (YOB=1960) |
| 5807 | DNA received as 1010045807 (YOB=1995) |
| Blank | no template control, water added instead of DNA |

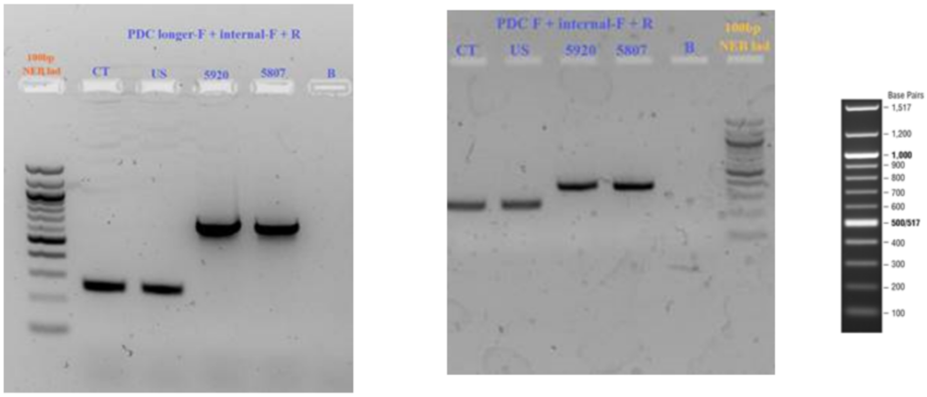


**Supplementary Figure 8: RT-PCR of *PDC* deletion.** Assay was designed with two sets of deletion spanning forward primers due to repetitive region near the 5’ end of *PDC.* For the deletion allele, higher bands (size 382 and 697) should be visible from both deletion spanning primers, with the 268bp band not amplifying due to loss of the complementary sequence inside the deletion. For the WT allele, the larger bands will not amplify while the 289bp band inside the deletion will be seen.

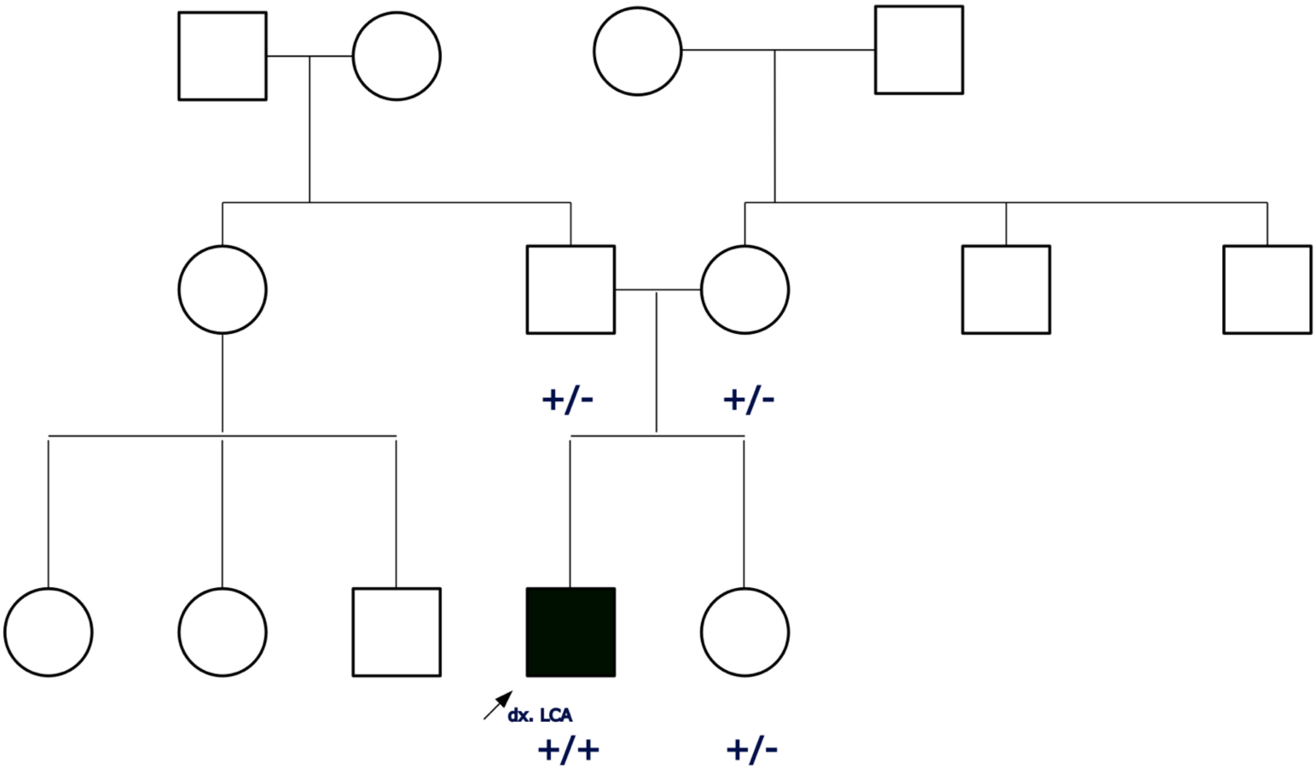


**Supplementary Figure 9:** ***PDC*** **pedigree.** Pedigree of *PDC* family where proband was diagnosed with Leber Congenital Amaurosis (LCA). Their sister is heterozygous for the deletion allele, along with both parents, none of which exhibit clinical signs of LCA.

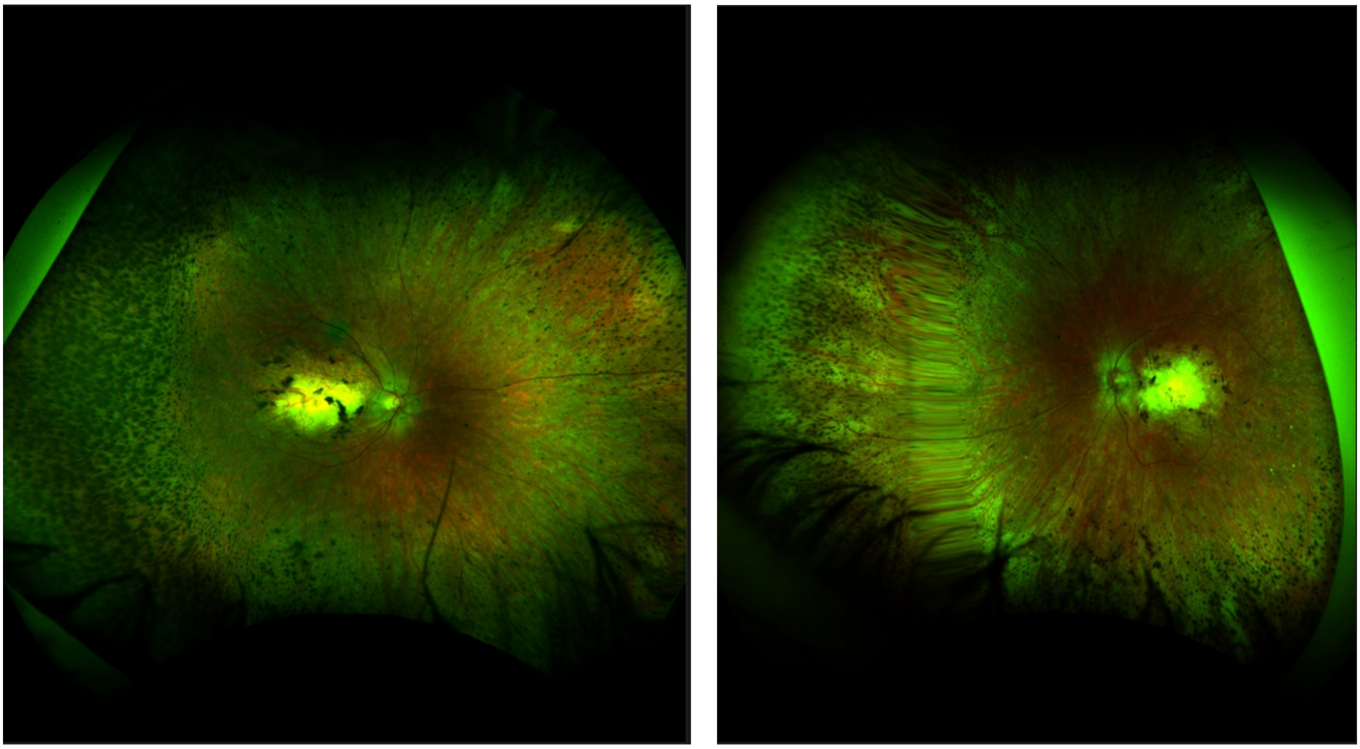

**Supplementary Figure 10: Fundoscopy of *PDC* proband.** Retinal imaging of proband from pedigree in Supplementary Figure 9.

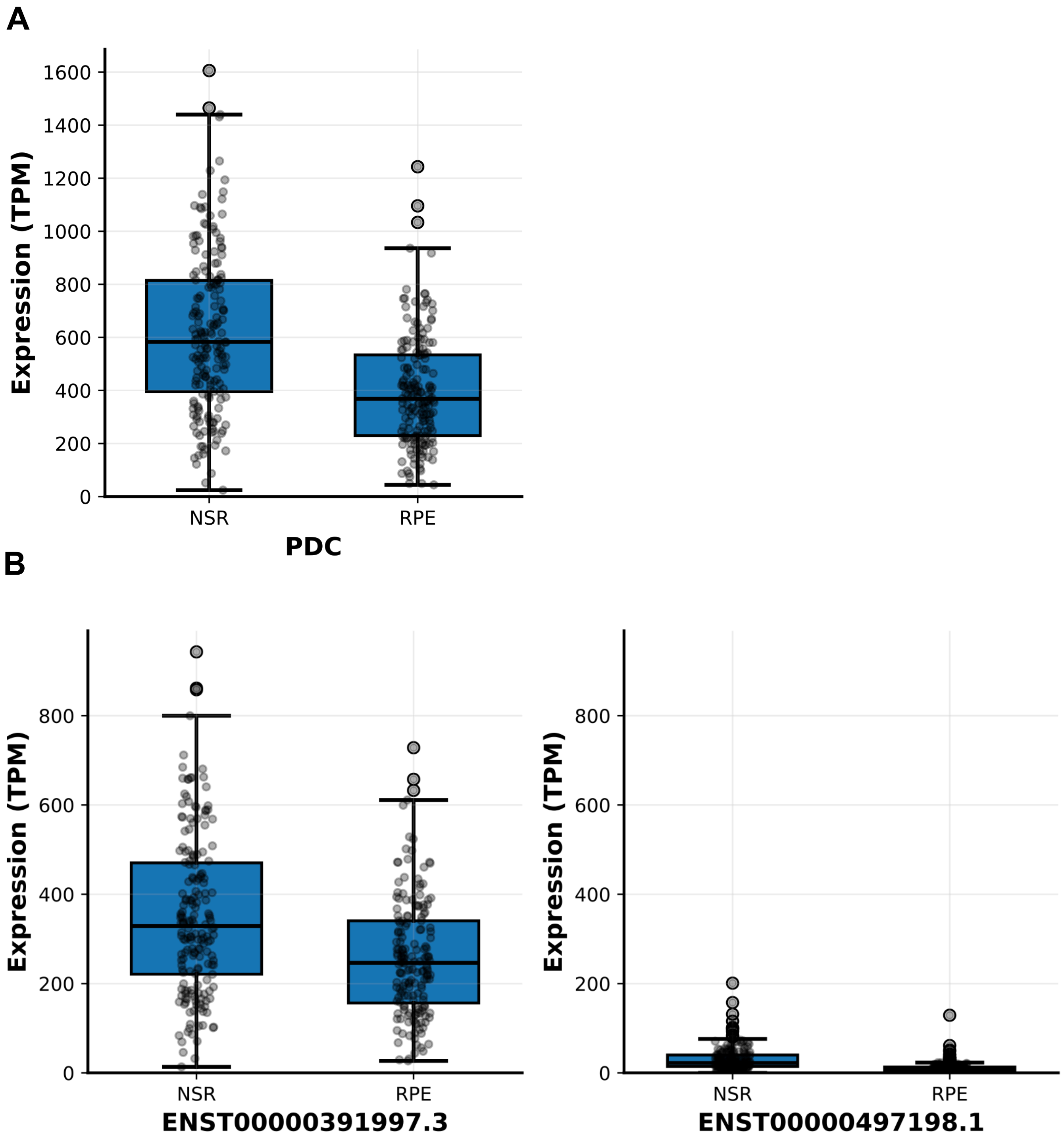


**Supplementary Figure 11: *PDC* retinal expression (A)** Gene wide expression of PDC in the neurosensory retina (NSR) vs the retinal pigment epithelium (RPE). **(B)** Transcript expression for the canonical transcript (ENST00000391997.3) vs an alternative isoform.

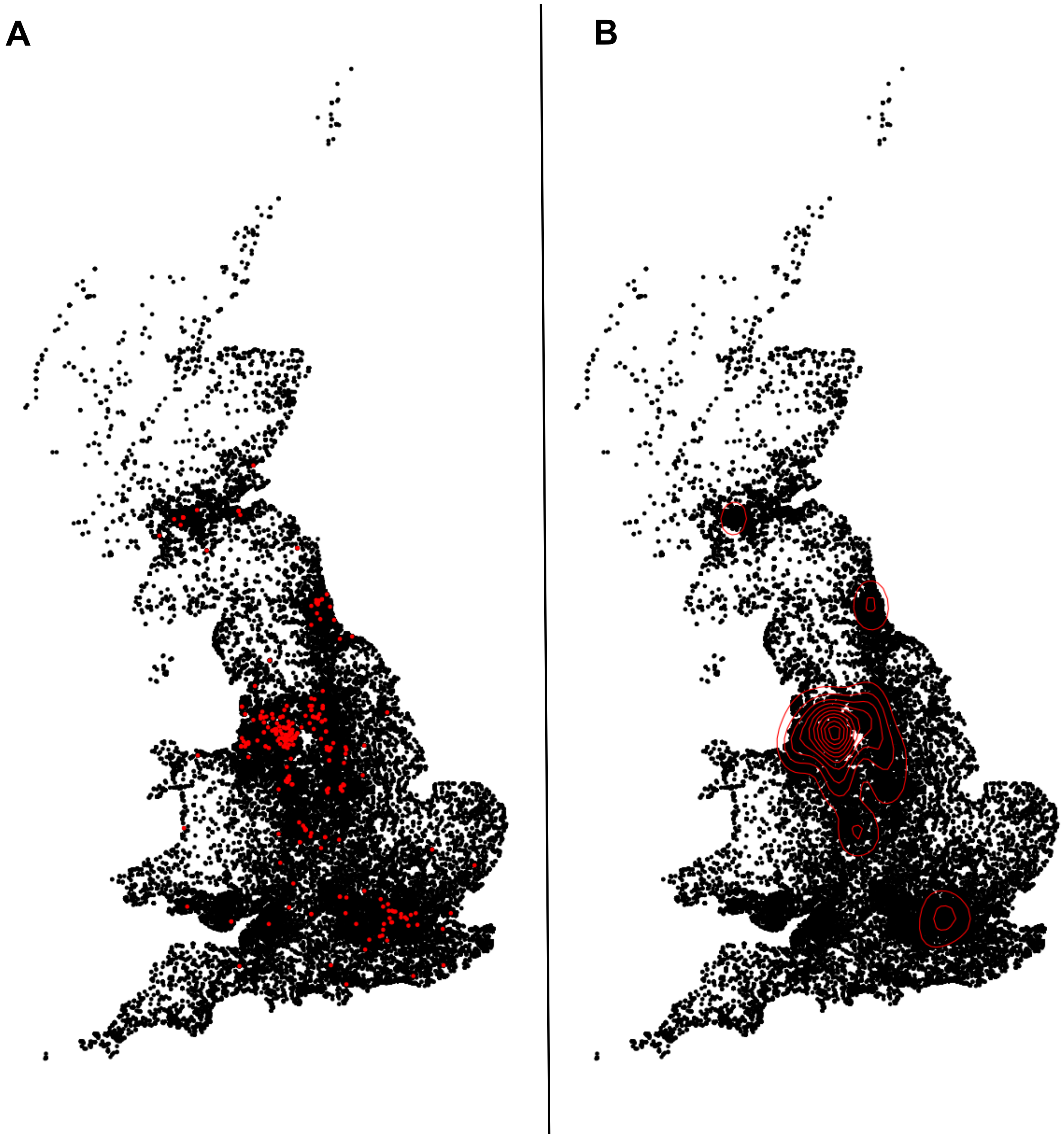


**Supplementary Figure 12: PDC carrier distribution across the UK (A)** Birth co-ordinates of *PDC* carriers seen in UK Biobank (n=311). **(B)** Density plot of the carrier co-ordinates.
